# Multi-omics biomarker selection and outlier detection across WHO glioma classifications via robust sparse multinomial regression

**DOI:** 10.1101/2024.08.26.24312601

**Authors:** João F. Carrilho, Roberta Coletti, Bruno M. Costa, Marta B. Lopes

## Abstract

Gliomas are aggressive brain tumors difficult to treat mostly due to their large molecular heterogeneity. This requires continuous improvement in the molecular characterization of the glioma types to identify potential therapeutic targets. Advances in glioma research are rapidly evolving, contributing to the updates of the WHO classification of tumors. Data analysis of multiple omics layers through classification and feature selection methods holds promise in identifying crucial molecular features for distinguishing between glioma types. We developed a robust and sparse classification workflow based on multinomial logistic regression to investigate the molecular landscape of gliomas. We considered transcriptomics and methylomics glioma profiles of patients labeled following the latest WHO glioma classification updates (2016 and 2021). Overall, our results show a notable improvement in glioma types separability for the 2021 WHO updated patient labels at both omics levels. Patients flagged as outliers for the 2016 WHO classification exhibited a molecular profile deviating from the one of the respective classes, which was more aligned with the current associated glioma type according to the 2021 WHO update. The methylomics profiles were particularly promising in the identification of outliers. These contributions will support further revisions of glioma molecular characterization and the development of novel targeted therapies.

## 1 Introduction

Gliomas are tumors that arise from the glial or progenitor cells of the Central Nervous System (CNS), being the main class of tumors of the brain and spinal cord. Over the past few decades, there has been extensive research in the field of biology focused on gliomas, which led to a rapidly evolving understanding of this class of tumors and histological subclasses at molecular levels. This evolving process has caused the World Health Organization (WHO) classification of gliomas to alter over time [1–3], as the advances in scientific research bring new intuitions into tumor inhabitation, ontogeny, and progression.

The status of isocitrate dehydrogenase (IDH) gene family, along with the combined loss of the short arm of chromosome 1 and the long arm of chromosome 19 (1p/19q codeletion), has been included into the guidelines for the WHO Classification of Tumors of the CNS in 2016 [2]. Consequently, astrocytoma and glioblastoma samples were further defined by their molecular characteristics, categorized as “IDH mutant” or “IDH wild-type”, coherently with the IDH status. Accordingly, the oligodendroglioma subtype was also molecularly defined, associating it with the joint presence of IDH mutation and 1p/19q co-deletion. Oligoastrocytoma samples according to the 2007 guidelines [1], i.e., gliomas characterized by mixed glial cells, were mostly reassigned to oligodendroglioma or astrocytoma, based on the status of the IDH mutation and presence or absence of 1p/19q co-deletion, respectively.

The latest WHO Classification of Tumors of the CNS, published in 2021 (2021-WHO) [3], emphasizes the relevance of molecular alterations in glioma classification. In adult-type gliomas, IDH testing is used as the separation factor between GBM and lower-grade glioma (LGG), namely astrocytoma and oligodendroglioma. Specifically, GBM is assigned in the presence of IDH wild-type coupled with certain molecular or histological features. On the other hand, if a sample exhibits IDH mutations, it is considered oligodendroglioma if chromosomes 1p and 19q are co-deleted, and astrocytoma otherwise.

The update of the classification guidelines is due to the advances in sequencing technologies that generate omics information. The implementation of machine learning algorithms to analyze omics data has revealed great potential, producing accurate results while effectively handling the molecular heterogeneity of tumors and the high dimensionality of omics data [4]. Feature selection techniques are essential for reducing the dataset dimension to a subset of representative features. This not only prevents overfitting but also aids in identifying relevant features related to tumor evolution. A state-of-the-art approach is feature selection via model regularization. This implies adding a penalty term in the model objective function which encourages sparsity in the model feature space. The elastic net regularizer [5] combines both *𝓁*_1_ (lasso) and *𝓁*_2_ (Ridge) regularization, being particularly suitable for omics studies where highdimensional datasets are generated and many irrelevant features might be present.

Classification tasks have the goal of estimating the sample category based on their characteristics and similarities with others assigned to the same label. Many examples demonstrate the great potentiality of such methods in the context of glioma, or cancer in general. Multinomial logistic regression (MLR) was applied to glioma clinical and single nucleotide polymorphism data from the Mayo Clinic to classify patients regarding molecular subtypes (IDH wild-type, IDH mutant 1p/19q non-codeleted and IDH mutant 1p/19q codeleted), returning relevant results to enhance understanding of gliomas [6]. Sparse MLR with the elastic net penalty (SMLR) was applied to breast cancer miRNA data to find biomarkers with great prognostic potential regarding breast cancer subtypes, achieving good accuracy results [7]. In the same study, six machine learning methods (MLR, MLR with the lasso or the ridge penalties, Support Vector Machines, and Random Forest) were applied to the reduced dataset composed of the biomarkers selected by SMLR for classification tasks, obtaining good classification accuracy values [7].

Biological datasets often contain outliers, observations that may stem from errors in sample labeling or atypical characteristics relative to their assigned group. Classical models may fail to identify outlying samples, leading to suboptimal solutions and reduced model accuracy. To mitigate variance caused by outliers, some authors suggest robust adaptations of statistical learning methods that allow the algorithm to handle and flag outlying samples. A common approach to robustness in linear and logistic regression models to high-dimensional datasets is the Least Trimmed Squares (LTS) estimator [8–10], which evaluates the error function at several subsets of samples and flags the ones out of the optimal subset as outliers. Segaert et al. [11] and Carrilho and Lopes [12] combined this estimator with the elastic net regularizer for outlier detection in triple-negative breast cancer and lower-grade glioma (LGG) RNA sequencing (RNA-seq) data from The Cancer Genome Atlas (TCGA), respectively, in binary classification scenarios via robust sparse binary logistic regression (rSLR). Robust SMLR with the LTS estimator (rSMLR) was recently proposed by Kurnaz and Filzmoser (2022) [13] for multi-class scenarios.

Despite the acknowledged importance of identifying outlier observations (e.g., patients, cells), uncovering their deviating origin in the feature space is crucial for gaining deeper insights into disease heterogeneity. A proper elementwise approach for outlier detection is expected to prevent discarding relevant information in the non-outlying data matrix elements [14]. The Detect Deviating Cells (DDC)^1^ algorithm [15] stands out as a pioneering elementwise outlier detection method for its ability to consider correlations between variables and deal with high-dimensional data. *Grosso modo*, this method flags the cells that stand out in each column in a standardized dataset and then for each cell, predicts its value according to the unflagged cells in its row whose columns are correlated with its column. A cell is then considered anomalous when its observed and predicted values differ significantly from each other. Segaert et al. [11] applied the DDC algorithm to breast cancer RNA-seq TCGA data, after robust feature selection by rSLR [10], for additional insights regarding the deviations of flagged patients in the selected features compared to the remaining observations in their respective classes.

In this study we explore the potential of sparse classification and outlier detection methods in identifying molecular biomarkers of glioma heterogeneity. The biological results of the application of these methods allow us to advance our understanding of the evolution of the classification of these tumors and ultimately enhance diagnostic therapeutic decision-making. Alongside disease stratification and biomarker selection, it is also essential to identify patients who exhibit deviations from the typical molecular pattern observed in patients of the same glioma type. Such advances perfectly align with current efforts to refine glioma classification. We applied multi-class classification with embedded feature selection to multi-omics glioma TCGA data with patient group labeling updated regarding the 2016 and 2021 WHO CNS classification guidelines. We developed SMLR and rSMLR models for both 2016-WHO [2] and 2021WHO [3] datasets to assess whether the 2021-WHO classification update enhanced the differentiation of glioma types and improved the allocation of outlier patients identified for the 2016-WHO classification. The classification task was complemented by visual inspection of the observations in a reduced dimensional feature space obtained via the Uniform Manifold Approximation and Projection (UMAP) algorithm [16], to understand the separability of the classes before and after feature selection. In the final phase of the analysis pipeline, we applied the DDC algorithm [15] to the 2016-WHO dataset, allowing the 2016 outliers’ molecular profile to be compared to the overall class pattern obtained by following the 2021 classification, on a featurewise basis.

## 2 Materials and Methods

### 2.1 Datasets

This study uses glioma RNA-seq and DNA methylation (DNA-meth) datasets from the TCGA portal. Data were extracted through the R functions `getFirehoseData`, available from the Bioconductor R package `RTCGA` [17], and `GDCquery`, `GDCdownload` and `GDCprepare`, from the Bioconductor R package `TCGAbiolinks` [18–20]. The glioma RNA-seq dataset contains the expression of 20501 genes from 659 patients. The glioma DNA-meth dataset includes the methylation level of 480457 sites from 653 patients. A pre-filtering procedure was applied to exclude non-valid entries, non-CpG sites, probes related to sexual chromosomes, cross-reactive and polymorphic CpGs [21–24], leading to 340427 CpG sites. The two datasets include sets of patients with significant overlap (80% shared samples).

In both omics datasets, three patient diagnostic labels are available, using the different editions of the WHO classification of tumors of the central nervous system: the original TCGA annotations, dated 2007 [1] (2007-WHO), and the updated diagnosis following the 2016 [2] (2016-WHO), and the 2021 [3] (2021-WHO) guidelines [25]. A cross-comparison between the diagnostic labels defined according to 2016-WHO (rows) and 2021-WHO (columns) in both (a) RNA-seq and (b) DNA-meth datasets can be found in Table 1. One can see that label changes occur mainly across the pairs GBM and astrocytoma (LGG-a), LGG-a and oligodendroglioma (LGG-od), and 75% previously unclassified samples in 2016-WHO became GBM in 2021-WHO.

**Table 1:**
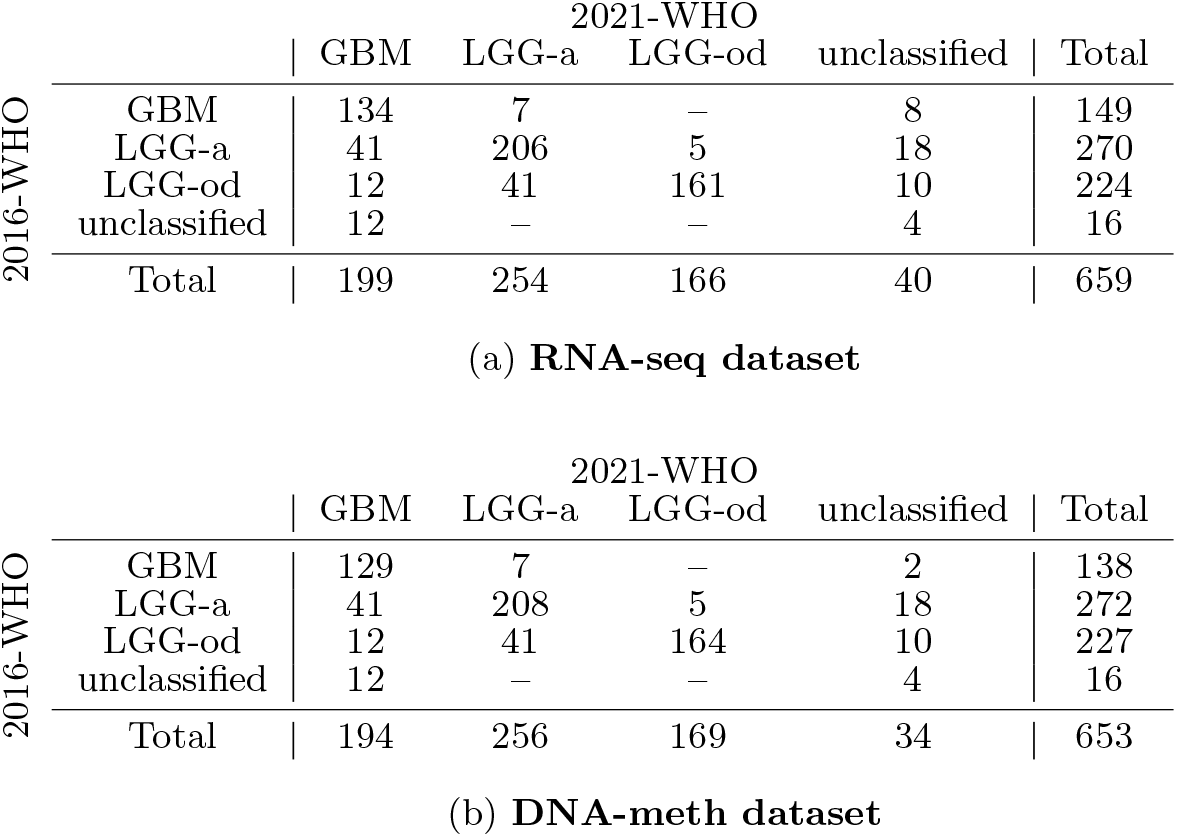
Diagnostic labels defined according to 2016-WHO (rows) and 2021-WHO (columns) in (a) RNA-seq, and (b) DNA-meth datasets. The numbers in each cell represent the number of patients that moved from the class (glioma type) in the rows to the class in the columns with the classification update (LGG-a, astrocytoma; LGG-od, oligodendroglioma).

### 2.2 Multinomial logistic regression with the elastic net penalty (SMLR)

Let us assume we have *N* observations ***x***_1_, …, ***x***_*N*_ of *p* variables. Each observation is also associated with a categorical factor variable *G*_*i*_ with *K* possible levels. The multinomial logistic regression estimates the probability of a given sample to be attributed to a level *𝓁* = 1, …, *K*, under the assumption 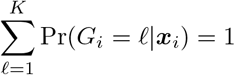 for each observation *i*.

This can be modeled as [26, 27]:

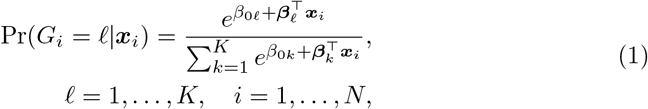

where ***β***_*𝓁*_ = (*β*_1*𝓁*_, …, *β*_*p𝓁*_) are coefficients estimated by minimizing the following negative log-likelihood function:

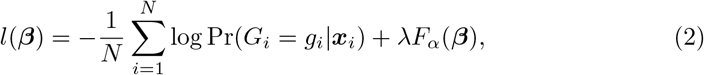

where ***β*** denotes the matrix of dimension (*p* + 1) × *K*, with the parameters *β*_0*𝓁*_ and ***β***_*𝓁*_ in its columns.

In equation (2), *g*_*i*_ represents the observed level of the variable *G*_*i*_ and the last term represents the penalty function, i.e., the term ensuring sparsity in the model, controlled by the regularization parameter *λ*. In this study, we used the Elastic net (EN) penalty [5], therefore:

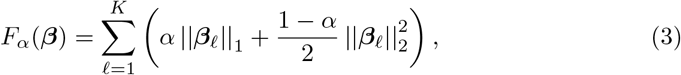

where the value of *α* ∈ [0, 1] regulates the balance between lasso and ridge penalties [5].

### 2.3 Robust multinomial logistic regression with the elastic net penalty and the least trimmed squares estimator (rSMLR)

SMLR is a powerful tool when performing feature selection on multinomial highdimensional data, however, outlying observations might increase the variance of the estimator. To address this issue, Kurnaz and Filzmoser proposed the robust SMLR (rSMLR) method [13], which identifies the observations that deviate from the rest of the data by integrating the LTS estimator [8] into the SMLR algorithm.

The subset *H* ⊆ {1, 2, …, *N*} and coefficients *β*_0*𝓁*_ and ***β***_*𝓁*_, *𝓁* = 1, …, *K* are then optimized by minimizing the penalized negative trimmed log-likelihood function in Equation (4), under the constraint |*H*| = *h*:

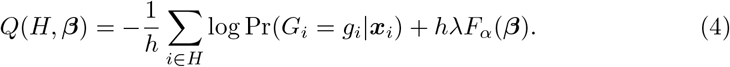

Parameters *α* and *λ* are optimized by 5-fold pairwise cross-validation from an input grid, resulting in the values *α*_opt_ and *λ*_opt_. Due to the computational cost, the rSMLR algorithm returns an approximate solution, from the convergence of an iterative method to minimize the function in Equation (4) and a further re-weighting step (Equation (5)) to increase the efficiency of the LTS estimator:

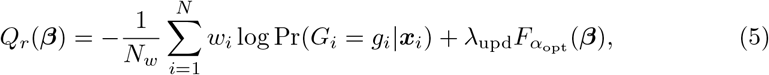

where *N*_*w*_ is the number of non-zero weights *w*_*i*_ and *λ*_upd_ is optimized by 5-fold crossvalidation for the previously obtained *α*_opt_ value. For further details, refer to Kurnaz and Filzmoser [13].

### 2.4 Detect Deviating Cells (DDC)

The DDC algorithm, proposed by Rousseeuw and Van Den Bossche (2018) [15] is a powerful tool for measuring each feature’s effect in observations flagged as outliers. Conversely to rSMLR, which detects outliers based on a rowwise approach, this method is able to estimate the influence of a single variable on all the patients, considering the dataset as a matrix composed by *N* × *p* entries we call cells.

By fixing an observation (row), DDC provides an estimate of the value of each data cell, based on those of the variables correlated with it, row. Then, the robustly standardized residual from the estimated and the actual cell value is computed, and the cell is flagged as an outlier if the corresponding residual’s absolute value exceeds 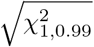 (i.e., the squared root of the 0.99-quantile of the *χ*^2^ distribution with 1 degree of freedom) [15].

### 2.5 Analysis pipeline

Figure 1 shows a schematic representation of the proposed multi-omics analysis. As part of data preprocessing, patients from both RNA-seq and DNA-meth datasets were associated with the diagnostic labels according to 2016-WHO and 2021-WHO [25]. Unclassified samples were not considered in our study. Before applying the classification algorithms, we split the datasets into training (75%) and test (25%) sets maintaining the original proportion of samples in each class. SMLR classification has been repeated for 100 runs, while rSMLR classification was applied over 30 runs due to the high computational cost of this method.

**Fig. 1:**
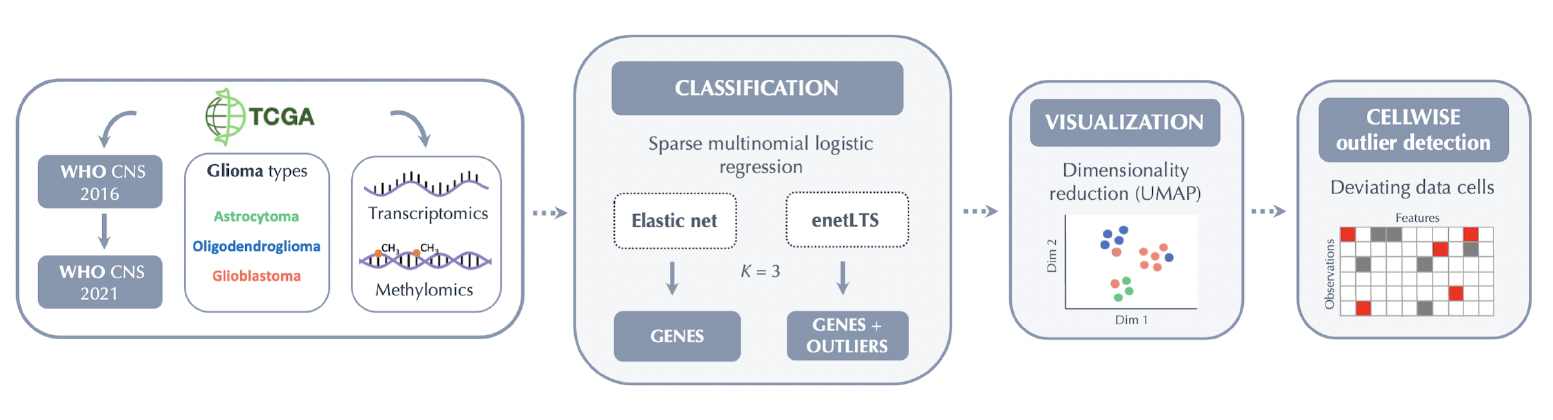
Overview of the analysis pipeline developed for glioma biomarker discovery and outlier detection.

The proposed methodology first applies classification to normalized (z-score) RNA-seq dataset, for both train and test sets. Features with a null standard deviation were discarded, leading to a total of 20200 variables. To overcome the problem of the high dimensionality of the DNA-meth dataset, it has been included only in the second stage of the analysis. Specifically, the number of DNA-meth variables was reduced to the subset of methylation sites associated with the genes previously selected by classification algorithms applied to RNA-seq data, with *α* = 0.1. The map linking genes and methylation sites was obtained by the R package `methylGSA` [28].

For both classification methods (SMLR and rSMLR), a sequence of *α* parameters was tested, from 0 (Ridge) to 1 (lasso), with a step of 0.1. The rSMLR model also requires an initial sequence of values for *λ*, set as {0.01, 0.02, …, 0.09, 0.1, 0.2, …, 0.9, 1}. *λ* and *α* were jointly estimated by pairwise cross-validation. Then, after that estimation, an updated value of *λ* was obtained in the re-weighting step. Classification by SMLR and rSMLR was performed using the R packages `glmnet` [27], and `enetLTS` [29], considering both the assigned patients’ labels (2016-WHO and 2021-WHO).

A feature was considered to be selected by a classification method when it was identified in at least 75 out of 100 train/test splits. Analogously, patients were considered misclassified if they were systematically assigned to the wrong class in at least 75% of the runs. A similar approach was considered for outlier identification, by taking into account that outliers can be detected only if they appear in the training set. Therefore, patients were defined as outliers if flagged by at least 75% of the training sets that include the sample.

After rSMLR classification, the DDC method was used by considering only the outliers detected from the 2016-WHO classification, to study their molecular profiles in light of the latest guideline release (2021-WHO). Every stage of our study was supported by UMAP visualization performed by `umap` R package [30] to facilitate the interpretation of the results. All procedures were implemented using version 4.1.3 of the `R` statistical software [31].

## 3 Results and Discussion

A prior graphical visualization of the data and the corresponding class membership in a subspace of reduced dimension was performed through the UMAP before the classification task. Figure 2 shows UMAP representations of the complete RNA-seq dataset, labeled according to 2016 and 2021 WHO classifications (left and right panels, respectively). It reveals a great improvement in patient groups’ homogeneity with the 2021 classification. For instance, in the left panel, we can observe a group of samples (bottom-left) including GBM and LGG, which in 2021 were mostly classified as GBM (right panel). The 2021-WHO also improves the distinction between the two LGG types (LGG-a and LGG-od), leading to more homogeneous groups. It can also be noted that unclassified samples are located near the GBM group on both left and right panels, which can be an indicator of the class they could potentially belong to. Unclassified samples were not considered in the subsequent analyses, as they were not included in the supervised analysis due to the lack of an updated diagnostic label.

**Fig. 2:**
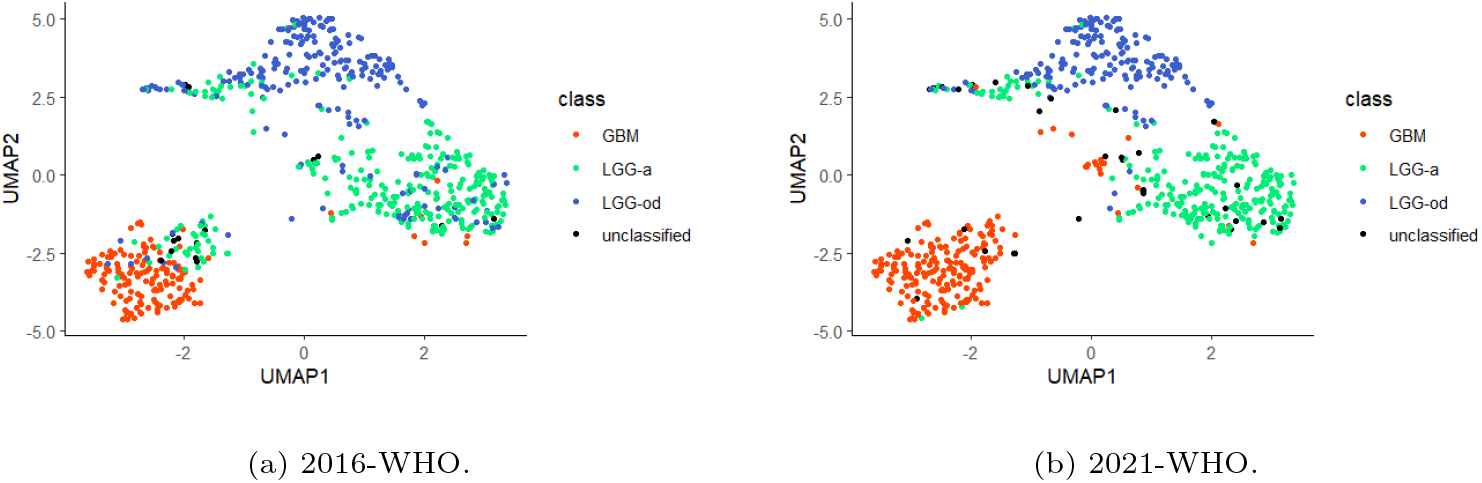
UMAP representations of the RNA-seq dataset. The two panels highlight with different colours the sample labels, assigned according to (a) 2016-WHO and (b) 2021-WHO. GBM: glioblastoma 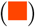. LGG-a: astrocytoma 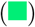. LGG-od: oligodendroglioma 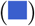. unclassified: unclassified patients 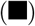.

Despite the pointed changes between classifications, both UMAP representations highlight the separability of glioma classes, which favors the application of sparse multinomial methods to identify the features that stand out as key diagnostic biomarkers in glioma-type classification. In the following, we show the results obtained by the application of SMLR and rSMLR models to the glioma omics data. We discuss the two model outcomes separately from a multi-omics perspective, according to the pipeline defined in Section 2.5.

### 3.1 SMLR models

SMLR was applied to RNA-seq data with patients classified according to both 2016-WHO and 2021-WHO. The overall SMLR performances by varying the parameter *α* are summarized in Table 2.

**Table 2:** Results obtained with SMLR models regarding the RNA-seq dataset. *β*_*ij*_ ≠ 0: number of coefficients which obtained a non-null value in at least 75% of the models; no. features: number of features corresponding to *β*_*ij*_ *≠* 0 coefficients; AUC: area under the ROC curve value; Sd: sample standard deviation for the associated value.

| $\alpha$ | $\beta_{ij} \neq 0$ (no. features) | | AUC Training Set | | | | AUC Test Set | | | |
| --- | --- | --- | --- | --- | --- | --- | --- | --- | --- | --- |
|  | 2016-WHO | 2021-WHO | 2016-WHO |  | 2021-WHO |  | 2016-WHO |  | 2021-WHO |  |
|  |  |  | Mean | Sd | Mean | Sd | Mean | Sd | Mean | Sd |
| 0 | 60579 (20193) | 60546 (20182) | 0.9486 | 0.0074 | 0.9961 | 0.0027 | 0.9337 | 0.0163 | 0.9879 | 0.0075 |
| 0.1 | 144 (143) | 299 (291) | 0.9506 | 0.0049 | 0.9947 | 0.0028 | 0.9488 | 0.0123 | 0.9906 | 0.0052 |
| 0.2 | 63 (63) | 149 (148) | 0.9513 | 0.0045 | 0.9947 | 0.0026 | 0.9495 | 0.0127 | 0.9908 | 0.0056 |
| 0.3 | 40 (40) | 93 (93) | 0.9521 | 0.0041 | 0.9942 | 0.0027 | 0.9504 | 0.0119 | 0.9906 | 0.0051 |
| 0.4 | 31 (31) | 57 (57) | 0.9525 | 0.0045 | 0.9940 | 0.0027 | 0.9510 | 0.0115 | 0.9909 | 0.0050 |
| 0.5 | 22 (22) | 42 (42) | 0.9526 | 0.0046 | 0.9941 | 0.0027 | 0.9508 | 0.0116 | 0.9911 | 0.0046 |
| 0.6 | 19 (19) | 36 (36) | 0.9529 | 0.0046 | 0.9941 | 0.0025 | 0.9505 | 0.0116 | 0.9911 | 0.0046 |
| 0.7 | 17 (17) | 29 (29) | 0.9524 | 0.0049 | 0.9942 | 0.0025 | 0.9497 | 0.0116 | 0.9912 | 0.0045 |
| 0.8 | 16 (16) | 23 (23) | 0.9524 | 0.0046 | 0.9944 | 0.0025 | 0.9498 | 0.0116 | 0.9914 | 0.0046 |
| 0.9 | 12 (12) | 19 (19) | 0.9525 | 0.0047 | 0.9949 | 0.0022 | 0.9500 | 0.0110 | 0.9916 | 0.0045 |
| 1 | 10 (10) | 17 (17) | 0.9526 | 0.0049 | 0.9953 | 0.0020 | 0.9501 | 0.0110 | 0.9915 | 0.0048 |

Following 2016-WHO, the SMLR models provided good performances, with mean multinomial area under the ROC curve (AUC) values [32, 33] of approximately 0.95 and low-expressed standard deviation values for all *α* values regarding both training and test sets. When considering 2021-WHO, the performances further improved, with an average AUC value around 0.99 for all models and small variances.

When applying SMLR to the reduced DNA-meth dataset, good AUC values were obtained for both training and test sets (Table 3). A slight decrease in the model performance with respect to the RNA-seq outcomes was verified for 2016-WHO. Conversely, 2021-WHO led to similar AUC values to the ones presented in Table 2.

**Table 3:** Results obtained with SMLR models from the DNA-meth dataset. *β*_*ij*_ ≠ 0: number of coefficients which obtained a non-null value in at least 75% of the models; no. features: number of features corresponding to *β*_*ij*_ ≠ 0 coefficients; AUC: area under the ROC curve value; Sd: sample standard deviation for the associated value.

| $\alpha$ | $\beta_{ij} \neq 0$ (no. features) | | AUC Training Set | | | | AUC Test Set | | | |
| --- | --- | --- | --- | --- | --- | --- | --- | --- | --- | --- |
|  | 2016-WHO | 2021-WHO | 2016-WHO |  | 2021-WHO |  | 2016-WHO |  | 2021-WHO |  |
|  |  |  | Mean | Sd | Mean | Sd | Mean | Sd | Mean | Sd |
| 0 | 5814 (1938) | 15441 (5147) | 0.9287 | 0.0113 | 0.9927 | 0.0033 | 0.9046 | 0.0188 | 0.9880 | 0.0069 |
| 0.1 | 191 (179) | 270 (264) | 0.9378 | 0.0142 | 0.9927 | 0.0026 | 0.9078 | 0.0201 | 0.9902 | 0.0065 |
| 0.2 | 103 (99) | 165 (163) | 0.9346 | 0.0156 | 0.9923 | 0.0023 | 0.9059 | 0.0197 | 0.9901 | 0.0063 |
| 0.3 | 70 (69) | 119 (118) | 0.9335 | 0.0146 | 0.9928 | 0.0023 | 0.9045 | 0.0196 | 0.9898 | 0.0059 |
| 0.4 | 54 (53) | 96 (95) | 0.9341 | 0.0149 | 0.9930 | 0.0024 | 0.9044 | 0.0191 | 0.9898 | 0.0058 |
| 0.5 | 47 (46) | 77 (76) | 0.9329 | 0.0135 | 0.9931 | 0.0024 | 0.9046 | 0.0190 | 0.9896 | 0.0056 |
| 0.6 | 42 (41) | 63 (62) | 0.9313 | 0.0120 | 0.9933 | 0.0024 | 0.9047 | 0.0201 | 0.9894 | 0.0058 |
| 0.7 | 33 (33) | 45 (44) | 0.9317 | 0.0134 | 0.9933 | 0.0025 | 0.9043 | 0.0201 | 0.9893 | 0.0061 |
| 0.8 | 29 (29) | 34 (33) | 0.9316 | 0.0137 | 0.9934 | 0.0025 | 0.9037 | 0.0195 | 0.9894 | 0.0061 |
| 0.9 | 27 (27) | 23 (22) | 0.9323 | 0.0139 | 0.9933 | 0.0025 | 0.9041 | 0.0208 | 0.9881 | 0.0071 |
| 1 | 21 (21) | 15 (14) | 0.9317 | 0.0130 | 0.9934 | 0.0025 | 0.9033 | 0.0211 | 0.9886 | 0.0068 |

These results further support the improvement of the newest classification guidelines, which appear to be more in accordance with patients’ molecular profiles. Notably, for both omics layers and guidelines (2016-WHO and 2021-WHO), no decrease in model predictive performance was observed with feature selection for increased *α* values, highlighting the ability of sparse models generated to select relevant variables able to discriminate between glioma types.

To further discuss the samples that were assigned to the wrong class (i.e., misclassified) by the SMLR model, we considered specific values of the regularization parameter *α*. These values were chosen to select an interpretable amount of features, which might be comparable between both classification guidelines for each omic.

For the RNA-seq data, we inspected the results obtained for *α* = 0.3, in the case of 2016-WHO, and *α* = 0.5, for 2021-WHO. Instead, for the DNA-meth data results, we chose *α* = 0.4 for 2016-WHO and 2021-WHO classification, respectively. We chose these *α* values since the respective models achieved a very satisfactory performance while selecting a relatively small yet biologically informative subset of genes. Table 4 resumes the patients that were misclassified according to 2016-WHO (rows) by the SMLR model based on both RNA-Seq and DNA-meth data. Most misclassified patients by SMLR viewed their label changed with the update to 2021-WHO [3, 25], suggesting great accordance between the 2021-WHO classification and both transcriptomics and methylomics profile.

**Table 4:** Label changes in the misclassified patients by SMLR applied to the RNA-seq (*α* = 0.3) and DNA-meth (*α* = 0.4) datasets, from 2016 to 2021 WHO classifications (the numbers in each cell represent the number of patients which moved from the class in the rows to the class in the columns with the classification update).

|  |  | Misclassifications |  |  |  |
| --- | --- | --- | --- | --- | --- |
|  | 2016\2021 | GBM | LGG-a | LGG-od | unclassified |
| RNA-seq | GBM | — | 1 | — | — |
|  | LGG-a | — | 4 | 4 | — |
|  | LGG-od | 12 | 33 | 1 | 6 |
| DNA-meth | GBM | — | 6 | — | — |
|  | LGG-a | 7 | 3 | 5 | 1 |
|  | LGG-od | 12 | 26 | — | 2 |

Figure 3 compares the Venn diagrams of intersections of misclassified patients across RNA-seq and DNA-meth datasets regarding both classifications. Based on the 2016-WHO guidelines (Figure 3a), despite most mislabeled samples being identified in common by the two omics layers, the SMLR model also detected some cases that do not match the associated class only according to one omics. This result points out that possibly mislabeled patients can have discordant transcriptomics and methylomics profiles, thus, fostering a multi-omics integrated analysis. When considering the 2021-WHO labels (Figure 3b) the number of patients assigned to a different class is highly reduced, corroborating the better performances of the model in this case.

**Fig. 3:**
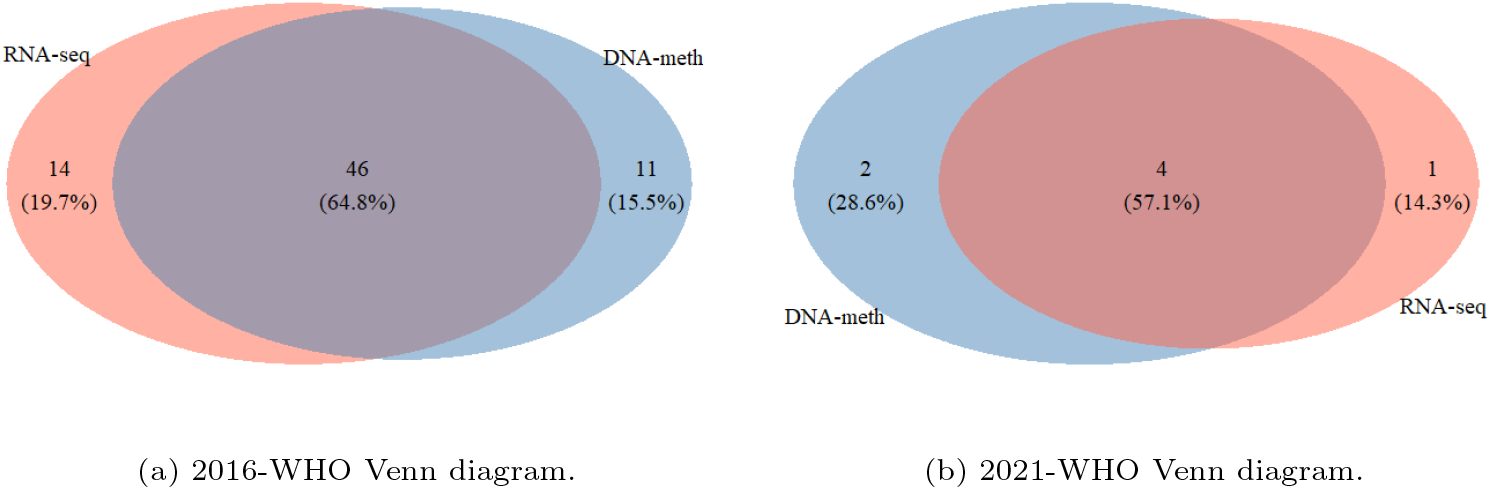
Venn diagrams with the intersection of the misclassified patients by SMLR-based classifiers applied to RNA-seq and DNA-meth datasets regarding (a) 2016-WHO and (b) 2021-WHO, where only patients comprised in both datasets were considered.

### 3.2 rSMLR models

Table 5 summarizes the results of rSMLR models. Notably, those performed a more relaxed feature selection for 2021-WHO than for 2016-WHO. Since the identified features correspond to the variables having non-null coefficients in at least 75% of the runs, this result suggests a high agreement in variable selection performed by the rSMLR models based on 2021-WHO. Also, this outcome is influenced by the regularization parameter choice, which the robust version of the model automatically estimates by pairwise cross-validation. When considering 2021-WHO classification, the parameter estimation were consistent among all the 30 runs, while for 2016-WHO different values were obtained. More details about the estimated regularization parameters are available in the Supplementary Material section (Table S1).

**Table 5:** Results obtained with rSMLR models (*β*_*ij*_ *≠* 0: number of coefficients which obtained a non-null value in at least 75% of the models; no. features: number of features corresponding to *β*_*ij*_ ≠ 0 coefficients; AUC: area under the ROC curve value; Sd: sample standard deviation for the associated value).

| Dataset | $\beta_{ij} \neq 0$ (no. features) | | AUC Training Set | | | | AUC Test Set | | | |
| --- | --- | --- | --- | --- | --- | --- | --- | --- | --- | --- |
|  | 2016-WHO | 2021-WHO | 2016-WHO<br>Mean | 2016-WHO<br>Sd | 2021-WHO<br>Mean | 2021-WHO<br>Sd | 2016-WHO<br>Mean | 2016-WHO<br>Sd | 2021-WHO<br>Mean | 2021-WHO<br>Sd |
| RNA-seq | 93 (91) | 670 (602) | 0.9389 | 0.0216 | 0.9959 | 0.0022 | 0.9340 | 0.0199 | 0.9908 | 0.0039 |
| DNA-meth | 535 (358) | 1557 (1331) | 0.8960 | 0.0070 | 0.9932 | 0.0011 | 0.8879 | 0.0194 | 0.9936 | 0.0034 |

The rSMLR based on 2016-WHO obtains slightly lower averaged AUC values when compared to SMLR. Nevertheless, also in the robust estimation, the model performances are remarkable, with an average AUC value around 0.93 (for RNA-seq) and 0.89 (for DNA-meth) in both training and test sets. Looking at the results based to 2021-WHO guidelines, the rSMLR model performs an accurate separation of the three defined glioma types, with high AUC values around 0.99 in train and test sets, for both omics layers.

Besides the variable selection based on the patients’ classes, the rSMLR method allows for the detection of outliers, i.e., patients who have a transcriptomics and/or methylomics profile differing from the one characterizing their class. To provide a visualization of the 2D-graphical distribution of the samples divided per class, and based on the subset of selected features in each case (2016-WHO and 2021-WHO) and omics, UMAP algorithm was applied to reduced datasets (Figure 4).

**Fig. 4:**
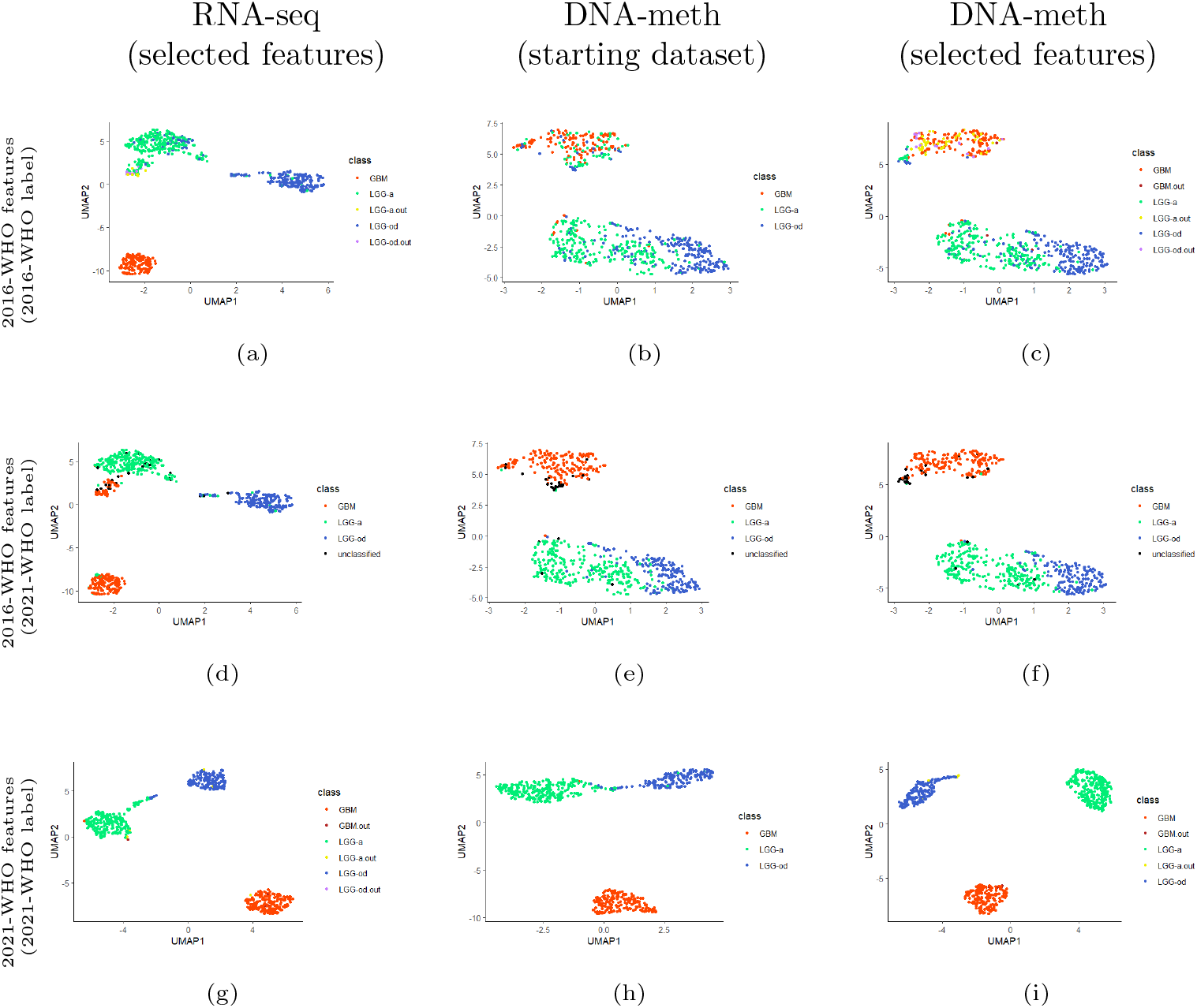
UMAP representations of patients based on the variables selected by the rSMLR models considering: (a-c) 2016-WHO guidelines, with colors assigned according to 2016-WHO; (d-f) 2016-WHO guidelines, with colors assigned according to 2021-WHO; (g-i) 2021-WHO guidelines, with colors assigned according to 2021-WHO. First column: selected features from RNA-seq dataset. Second column: starting DNA-meth dataset. Third column: selected features from DNA-meth dataset. GBM: patients with glioblastoma 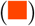. GBM.out: outlier patients within the glioblastoma class 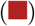. LGG-a: patients with astrocytoma 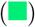. LGG-a.out: outlier patients within the astrocytoma class 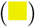. LGG-od: patients with oligodendroglioma 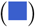. LGG-od.out: outlier patients within the oligodendroglioma class 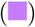. unclassified: unclassified patients 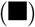.

The aim was to observe how the projection of the patients in a lower dimentional space changes, by considering only variables selected by the applied method, across the two omics. In particular, each line of the Figure 4 includes, in sequence, the UMAP graphs based on: the selected genes (rSMLR outcome based on RNA-seq data), the associated CpG sites (starting DNA-meth data), and the selected CpG sites (rSMLR outcome based on DNA-meth data). The first (Figures 4a to 4c) and last (Figures 4g to 4i) lines consider the subset of variables depending on the 2016-WHO and 2021-WHO labels, respectively. The samples (points) are colored according to their diagnostic labels. Instead, the line in the middle (Figures 4d to 4f), reports the same projection based on the 2016-WHO, while the colors are assigned based on the 2021-WHO classes, allowing an easier observation of the diagnostic label evolution. In each graph, the outlier samples are colored differently, based on the class to which they belong.

Overall, all UMAPs show a good separation of the three glioma types. However, when considering 2016-WHO labels, an overlap of the two LGG types is visible from RNA-seq data (Figure 4a). This class heterogeneity is further emphasized when moving to the DNA-meth layers (Figures 4b and 4c), where the groups appear highly mixed, with many LGG-a and LGG-od samples projected in the GBM area, most of which identified as outliers. Interestingly, if these samples are colored according to the 2021-WHO diagnostic labels, most of them change to GBM (Figure 4f), supporting the great improvement of the latest guidelines. This result is confirmed by the UMAPs obtained from the subset of features selected by the model considering the 2021-WHO labels, in which a nearly-perfect separation of the three types is obtained after the application of both steps of our pipeline (Figure 4i).

A quantification of outliers and misclassified samples resulting from rSMLR model based on 2016-WHO guidelines, with respect to their updated 2021-WHO diagnostic labels, is summarized in Table 6. As previously observed, most of the LGG outliers changed their labels to GBM, which fosters a more detailed cellwise study of these samples to understand the aspects that led to their identification. On the other hand, from the misclassification results, many LGG-od samples wrongly assigned can be noted, most of which would be classified as LGG-a following 2021-WHO guidelines. By focusing on DNA-meth data, the number of outliers and misclassified samples increases, and also some GBM cases appear. The majority of these additional samples, which were not pointed out by the analysis on RNA-seq data, would change their diagnostic label with the 2021-WHO guidelines update, suggesting a bigger discrepancy between samples profiles and glioma types in DNA-methylation than in gene expression. Figure 5 shows the Venn diagrams of outlier patients identified across RNA-seq and DNA-meth datasets regarding both classifications. Conversely to what observed for samples misclassified by SMLR model, most outliers were identified from methylomics, further supporting the relevance of this layer.

**Table 6:**
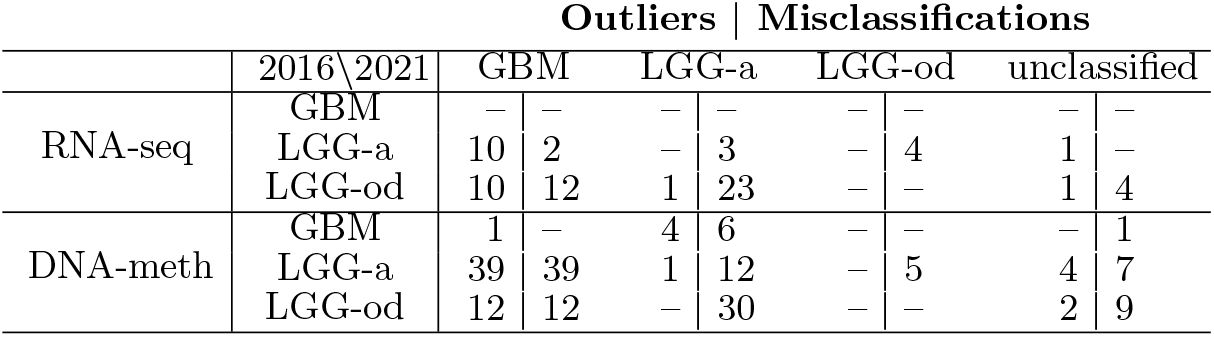
Patients detected as outliers or misclassified by the rSMLR model based on 2016-WHO, applied to the RNA-seq and DNA-meth datasets. The rows report the 2016-WHO labels, while the columns refer to their 2021-WHO update. Each cell represents the number of patients who changed their diagnosis with the 2021-WHO update, from the class in the rows to the class in the columns.

|  |  | Outliers \| Misclassifications |  |  |  |  |  |  |  |
| --- | --- | --- | --- | --- | --- | --- | --- | --- | --- |
|  | 2016\2021 | GBM |  | LGG-a |  | LGG-od |  | unclassified |  |
| RNA-seq | GBM | — | — | — | — | — | — | — | — |
|  | LGG-a | 10 | 2 | — | 3 | — | 4 | 1 | — |
|  | LGG-od | 10 | 12 | 1 | 23 | — | — | 1 | 4 |
| DNA-meth | GBM | 1 | — | 4 | 6 | — | — | — | 1 |
|  | LGG-a | 39 | 39 | 1 | 12 | — | 5 | 4 | 7 |
|  | LGG-od | 12 | 12 | — | 30 | — | — | 2 | 9 |

**Fig. 5:**
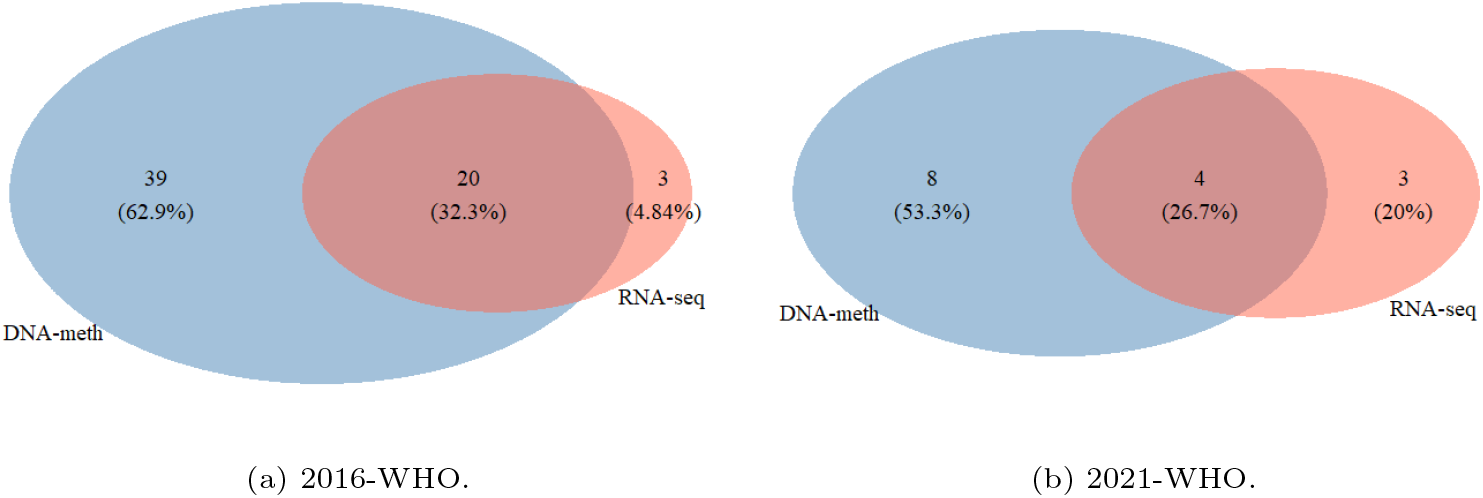
Venn diagrams with intersections of patients flagged by the rSMLR model as outliers from RNA-seq and DNA-meth datasets based on (a) 2016-WHO and (b) 2021-WHO. Only patients comprised in both omics datasets were considered.

The comparison between outliers and misclassified samples, depending on the guidelines and the omics layer considered, is visible in the Venn diagrams in Figure 6. Overall, an agreement between the two groups can be observed (i.e., outliers were mainly misclassified). Additionally, the models based on 2016-WHO labels led to a higher number of misclassified samples, confirming the difficulty in identifying the glioma types defined by 2016-WHO based on the patients’ molecular profiles.

**Fig. 6:**
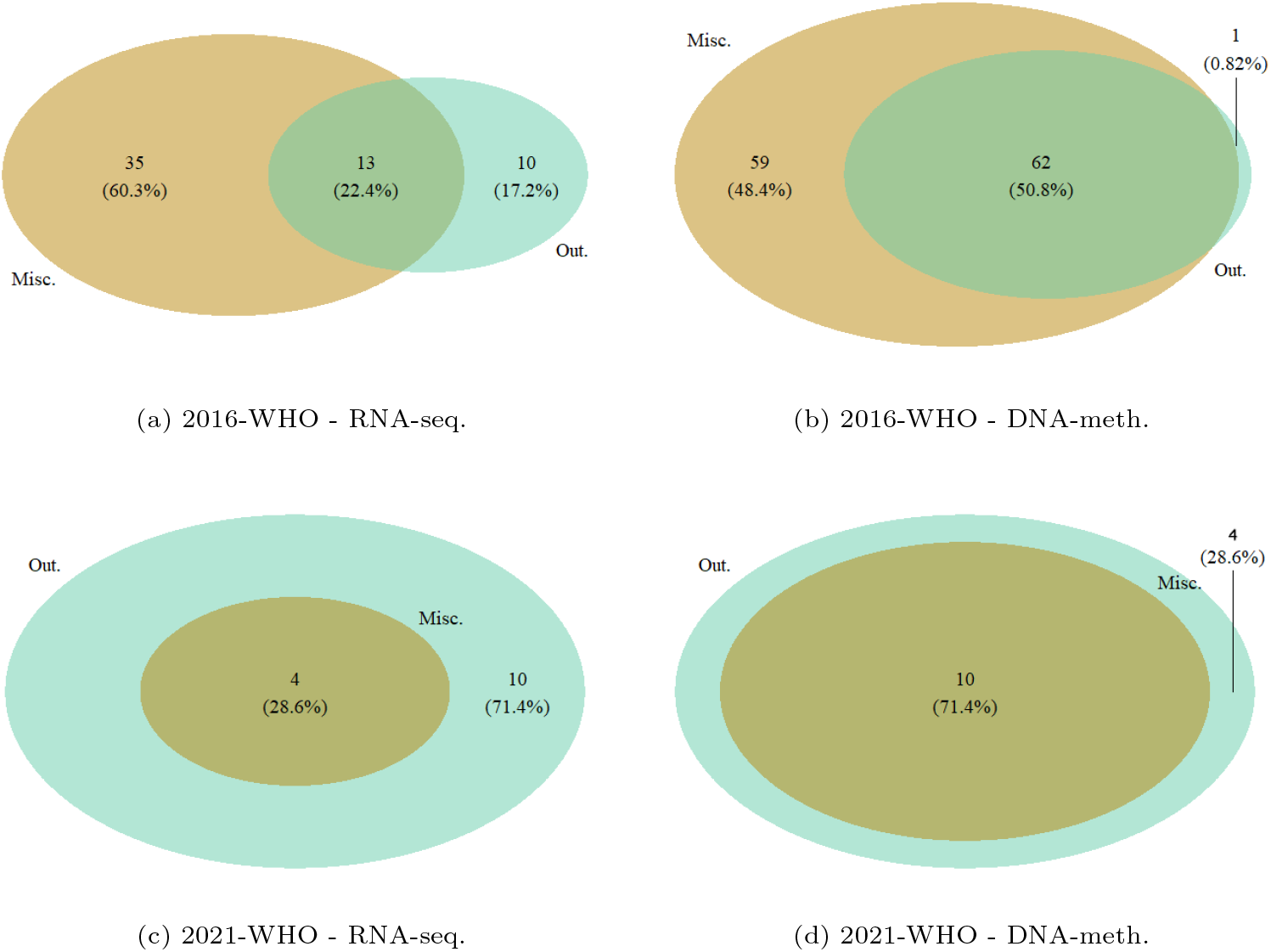
Venn diagrams with intersections of patients that were misclassified (Misc.) and flagged as outliers (Out.) by rSMLR models based on 2016-WHO (a-b) and 2021-WHO (c-d).

To analyze the molecular profile of the samples identified as outliers based on the 2016-WHO labels, the DDC algorithm was applied. Figure 7 illustrates the DDC results corresponding to RNA-seq data. Rows report all the outliers, together with a random subset of samples per type. To facilitate the comparison among patients’ profiles, outliers are sorted to be close to the samples belonging to the glioma type to which these would be associated according to the 2021-WHO. Columns list the variables that were selected by rSMLR model, grouped according to the glioma type to which they are related. Colored cells highlight the genes of a given sample having a different expression/methylation level compared to their predicted expected value (purple to blue tones for lower than expected, orange to red tones for higher than expected). By looking at Figure 7, based on RNA-seq data, it is possible to detect some common patterns between patients with different diagnoses. For instance, the first outliers’ group, coherently with GBM samples, presents a high deviation of the gene *RAB42* that is not common neither in LGG-a, nor in LGG-od patient profiles. Accordingly, the only LGG-od outlier sample changing the diagnosis to LGG-a based on 2021-WHO shows high expression of *C15orf21*, a gene the model detects to characterize LGG-a.

**Fig. 7:**
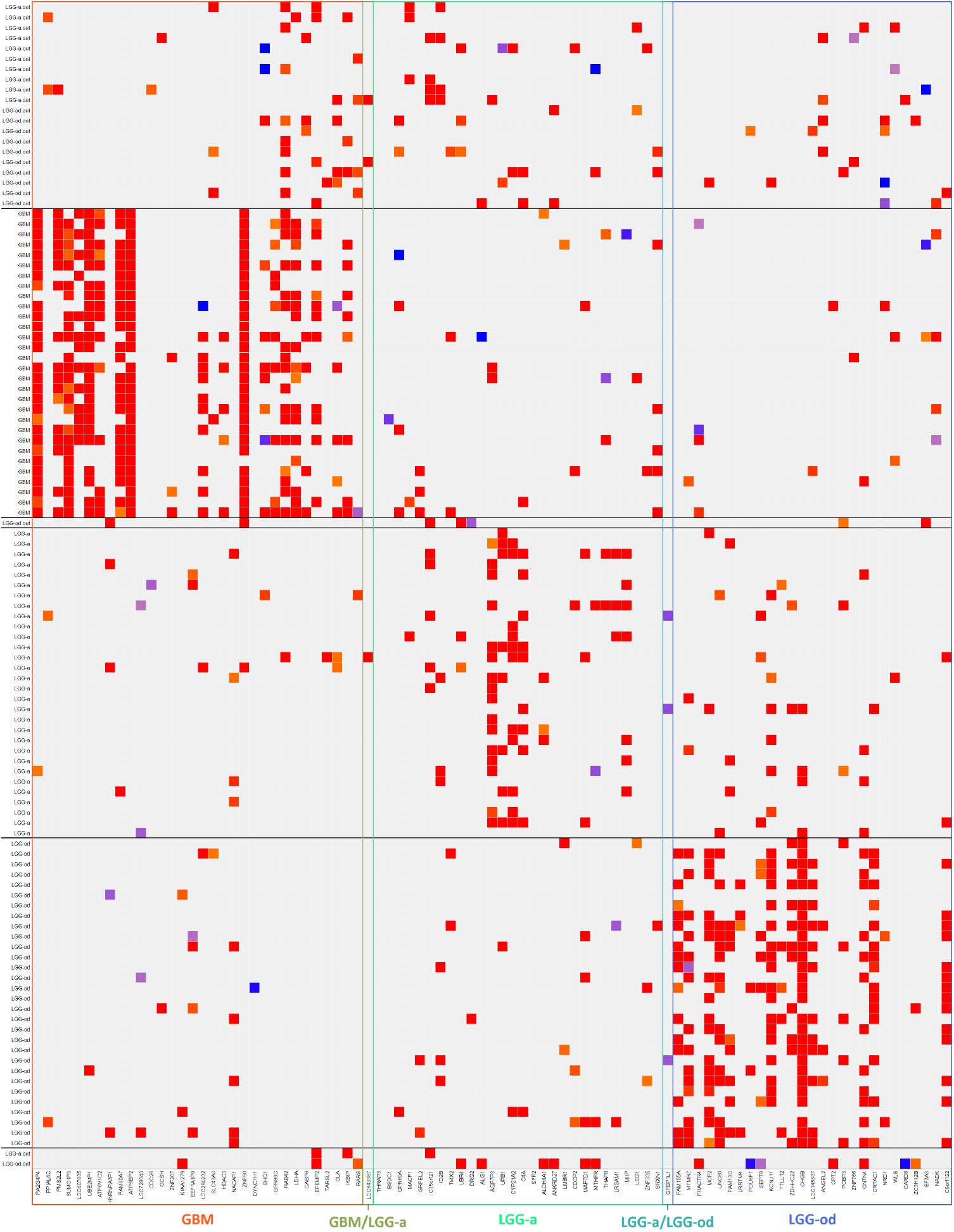
DDC plot for the features selected by the model from the RNA-seq dataset. Grey cells represent cells whose observed values do not deviate significantly from the expected value. Purple-to blue-colored cells represent observed values lower than their expected value while orange- to red-colored cells represent values higher than their expected value.

The DDC plots regarding the DNA-meth data are showed in the Supplementary Material (Figures S1 to S3). Due to the huge number of features selected in this case, each figure refer to the CpG sites selected for a given glioma type. From this representation, the pattern characterizing LGG-a and LGG-od samples is even more evident, and it highly differs from the GBM one. Moreover, the outliers’ methylomics profile matches with the one of their corresponding class based on 2021-WHO labels, suggesting a strong alignment between methylomics data and glioma types (see, as an example, the GBM outliers that would change to LGG-a with the 2021-WHO update). The DDC algorithm was also applied to further investigate outliers detected by considering the updated 2021-WHO guidelines for glioma classification. The corresponding Figures are provided in the Supplementary Material section (S4 to S9).

The top 20 features associated with each glioma type for RNA-seq dataset were further investigated across the literature. The relevant findings of this research are summarized in Table S2 (Supplementary Material), highlighting the genes’ influence on glioma or cancer processes. Table S3 (Supplementary Material) lists the top 20 features selected for DNA-meth dataset. This table reports methylation sites and their associated genes, together with their rankings in both methylomics and transcriptomics for each glioma type based on the rSMLR outcome.

## 4 Conclusion

The objective of this study was to identify molecular biomarkers of glioma heterogeneity, as well as the identification of outlying patients through their molecular profiles. This contributes to enhancing the molecular understanding of these tumors and support clinical and therapeutic decisions.

SMLR and rSMLR models were applied to glioma RNA-seq and DNA-meth data. Patient labeling according to the 2021-WHO notably improved model performances, based on both transcriptomics and methylomics data, with the outliers identified by rSMLR applied to 2016-WHO data corroborating the 2021-WHO patient labeling update. SMLR and rSMLR models also selected groups of transcriptomics and methylomics variables as potential diagnostic biomarkers, which might uncover relevant insights on glioma heterogeneity from a biological perspective. Most top-selected genes were already reported in the literature as being connected with glioma or cancer in general, therefore supporting the methodology used and fostering the biological validation of the non-reported genes.

Despite the outstanding performance of SMLR and rSMLR models when applied to 2021-WHO, the robust model still identified outliers (i.e. deviating molecular profiles) among the classes, which means that the supervised approach followed does not cover all glioma heterogeneity.

The results obtained by the models and the analysis of the outlying patients highlighted an important role of methylomics in outlier detection, suggesting that future updates of glioma classifications must focus on patient methylomics profiles as already pointed out in prior studies [34–36].

It will also be advantageous in future studies to use unsupervised methods, namely robust clustering, to investigate the within-group variability and identify new glioma subgroups towards the development of more personalized therapies.

## Data Availability

The updated diagnostic labels of the samples of the
TCGA glioma dataset used in this study are available
at: https://github.com/RobertaColetti/
Update-TCGA-glioma-WHOclassification.

https://github.com/RobertaColetti/Update-TCGA-glioma-WHOclassification

## Acknowledgements

This work was supported by national funds through Fundação para a Ciência e a Tecnologia (FCT I.P.) with references CEECIN-ST/00042/2021, UIDB/00297/2020 and UIDP/00297/2020 (NOVA MATH), UIDB/00667/2020 and UIDP/00667/2020 (UNIDEMI), and developed within the project “MONET: Multi-omic networks in gliomas” (PTDC/CCI-BIO/4180/2020). The results here obtained are based upon data generated by the TCGA Research Network: https://www.cancer.gov/tcga. We thank Professor Susan P. Holmes for her valuable revision and comments on the manuscript.

## Footnotes

1 *x*_*ij*_ elements of a data matrix *X*. Not to be interpreted in the biological sense.

