## Supplementary Material for "Multi-omics biomarker selection and outlier detection across WHO glioma classifications via robust sparse multinomial regression"

### S1 $(\alpha, \lambda)$ values selected by rSMLR for 2016-WHO

Table S1: Pairwise choices for $\alpha$ and $\lambda$ coefficients in each rSMLR run for 2016-WHO.

| $\alpha \backslash \lambda$ | 0.01 | 0.02 | 0.03 |
|---|---|---|---|
| 0.1 | 9 | 1 | 3 |
| 0.2 | 5 | 2 | – |
| 0.4 | 3 | – | – |
| 0.5 | 3 | – | – |
| 0.6 | 2 | – | – |
| 0.7 | 1 | – | – |
| 0.8 | 1 | – | – |

### S2 DDC plots

Pages s2-s10 contain the outputs of the application of the DDC algorithm to the subsets of genes and methylation sites which were selected by rSMLR models for 2016-WHO and 2021-WHO, where lines represent patients and columns represent features, organized by the associated class (or group of classes in the case features were associated with more than one class).

The black lines in each Figure separate rows of patients according to their classification and to their identification (or non-identification) as outliers.

The structure in row sets in subsection S2.1 (DDC plots regarding 2016-WHO) is as follows:

1. Outlying patients whose classification according to 2021-WHO is GBM;
2. A random sample of 30 non-outlying GBM patients;
3. Outlying patients whose classification according to 2021-WHO is LGGa;
4. A random sample of 30 non-outlying LGG-a patients;
5. A random sample of 30 non-outlying LGG-od patients;
6. Outlying patients without classification according to 2021-WHO.

This structure changes slightly in Section S2.2 (DDC plots regarding 2021-WHO). It becomes as follows:

1. Outlying patients in the GBM class;
2. A random sample of 30 non-outlying GBM patients;
3. Outlying patients in the LGG-a class;
4. A random sample of 30 non-outlying LGG-a patients;
5. Outlying patients in the LGG-od class[1];
6. A random sample of 30 non-outlying LGG-od patients.

Note that Figures S1 and S4 to S9 are rotated due to their width being larger than their height.

It is also relevant to point out that the random samples from each class are kept the same for each omics-guidelines combination.

[1]This set exists only in plots regarding rSMLR models applied to the RNA-seq dataset.

#### S2.1 DDC plots regarding 2016-WHO

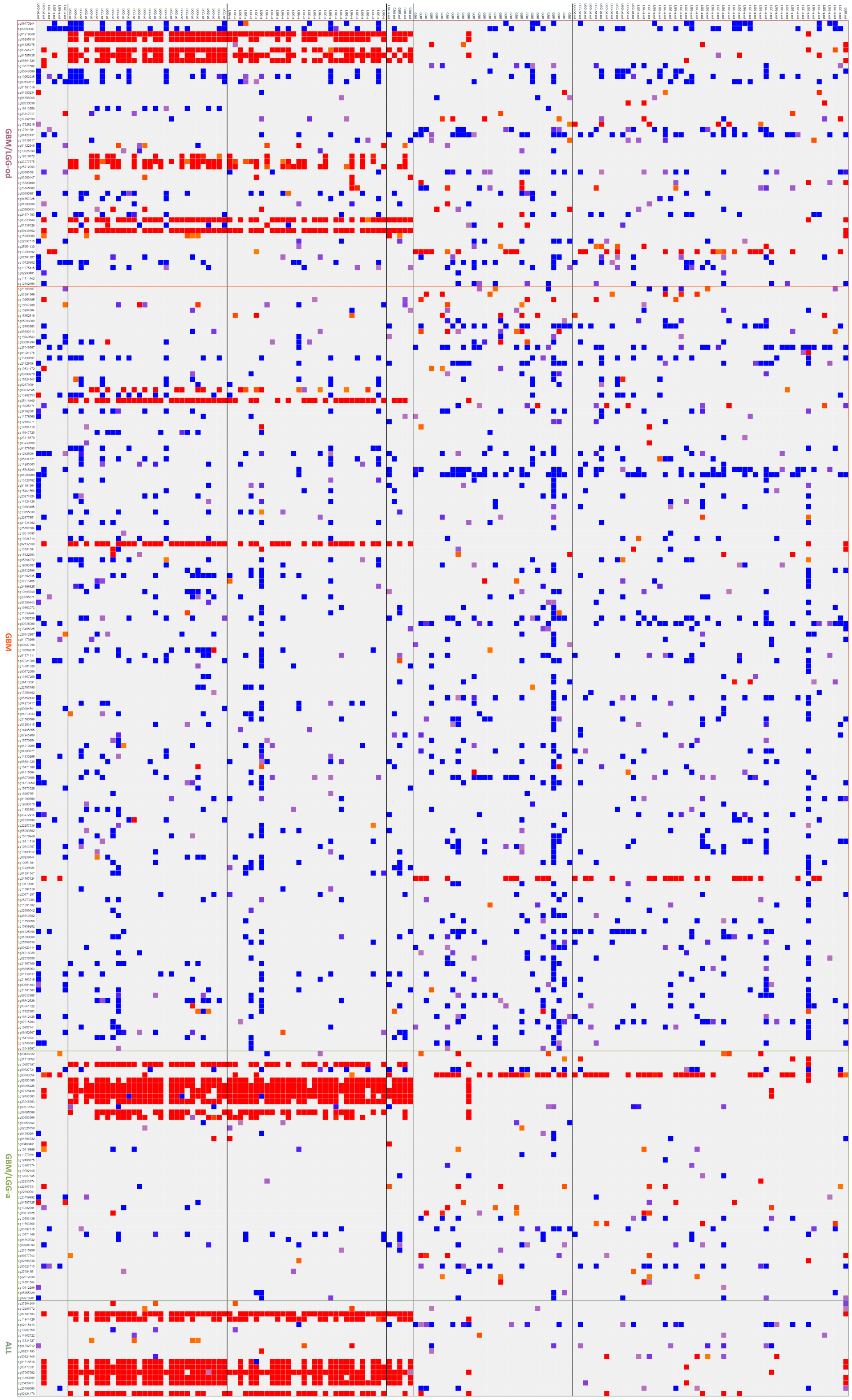


Figure S1: DDC plot for the features associated with GBM by the model from the DNA-meth dataset.

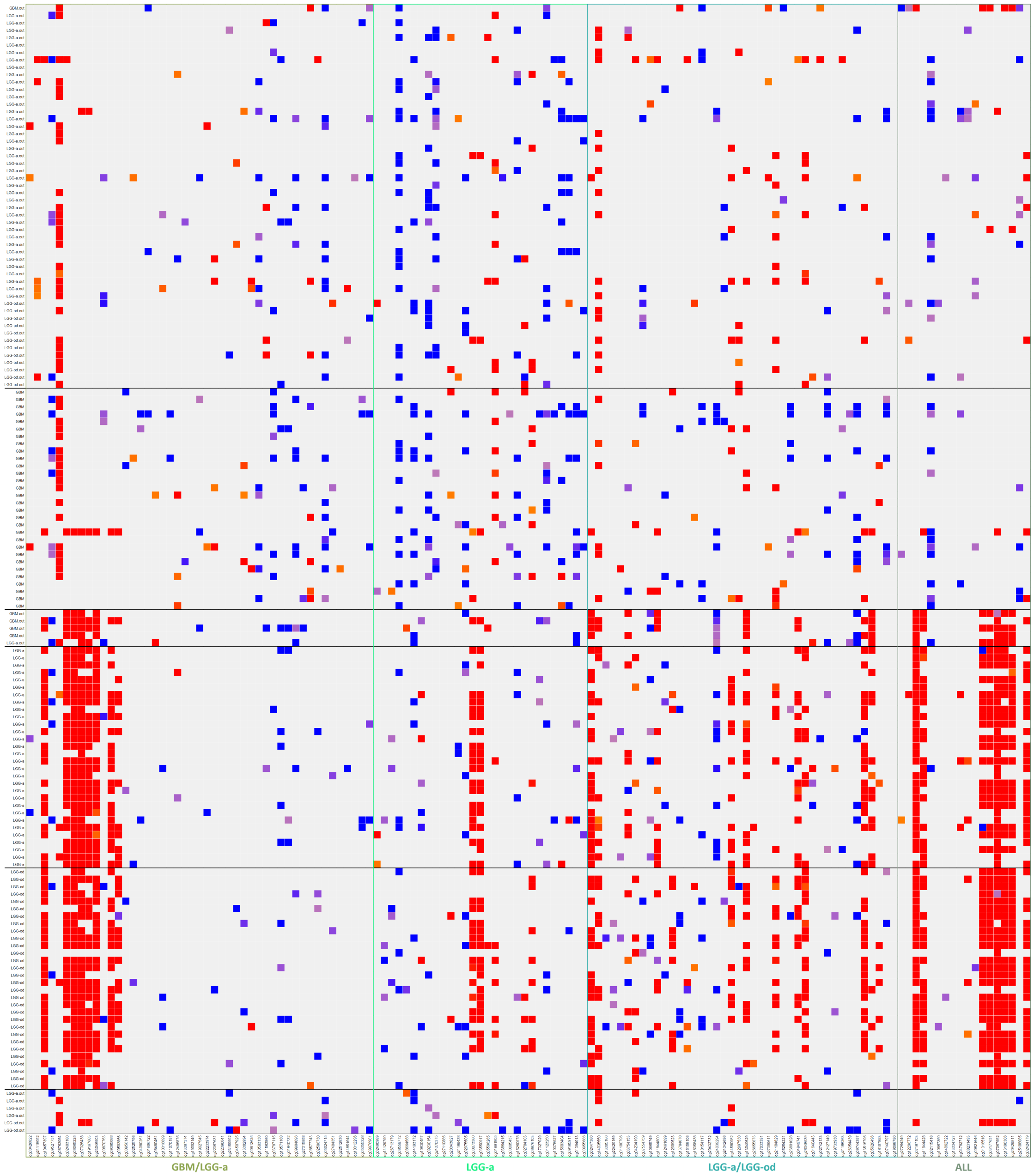


Figure S2: DDC plot for the features associated with LGG-a by the model from the DNA-meth dataset.

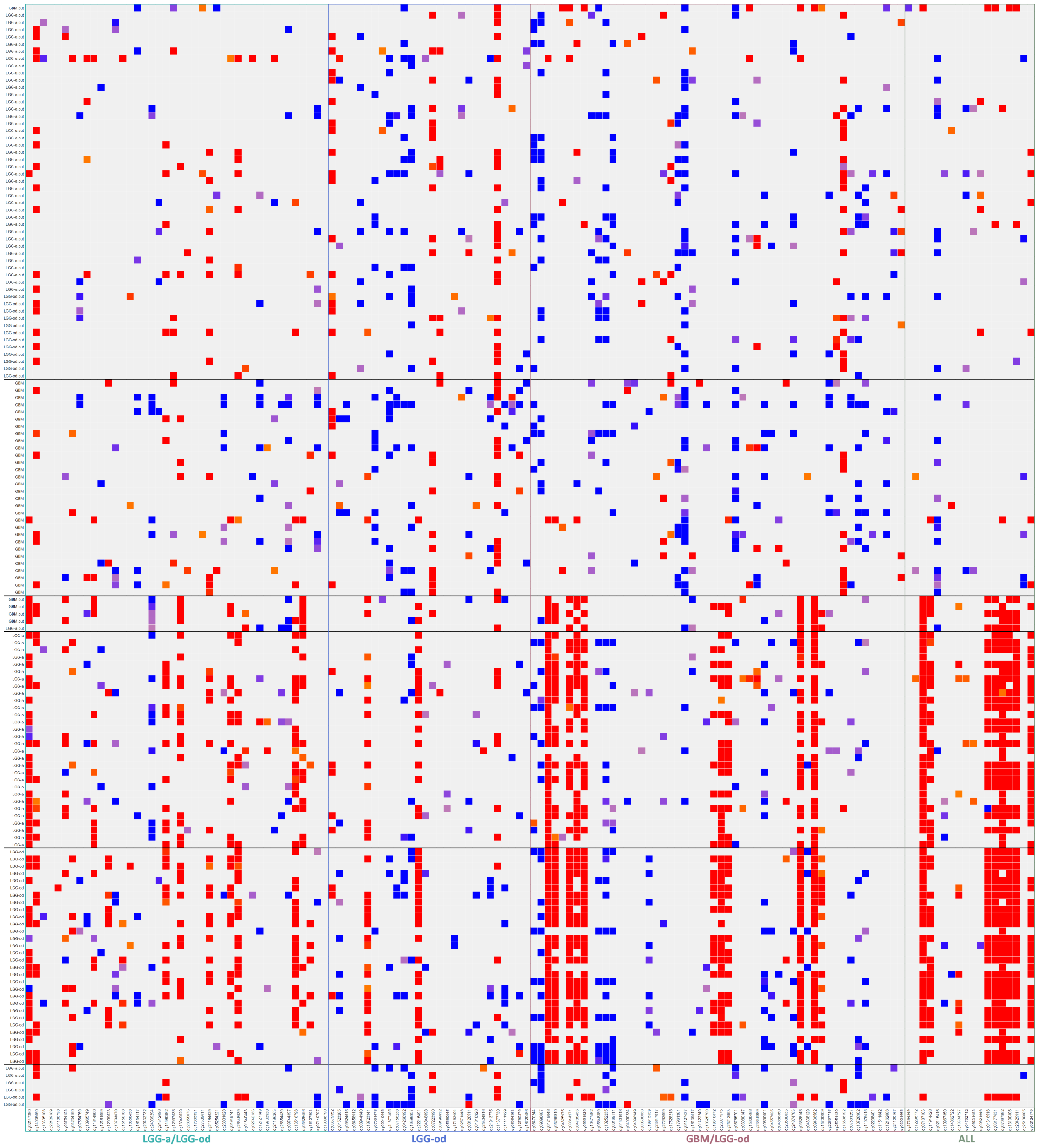


Figure S3: DDC plot for the features associated with LGG-od by the model from the DNA-meth dataset.

#### S2.2 DDC plots regarding 2021-WHO

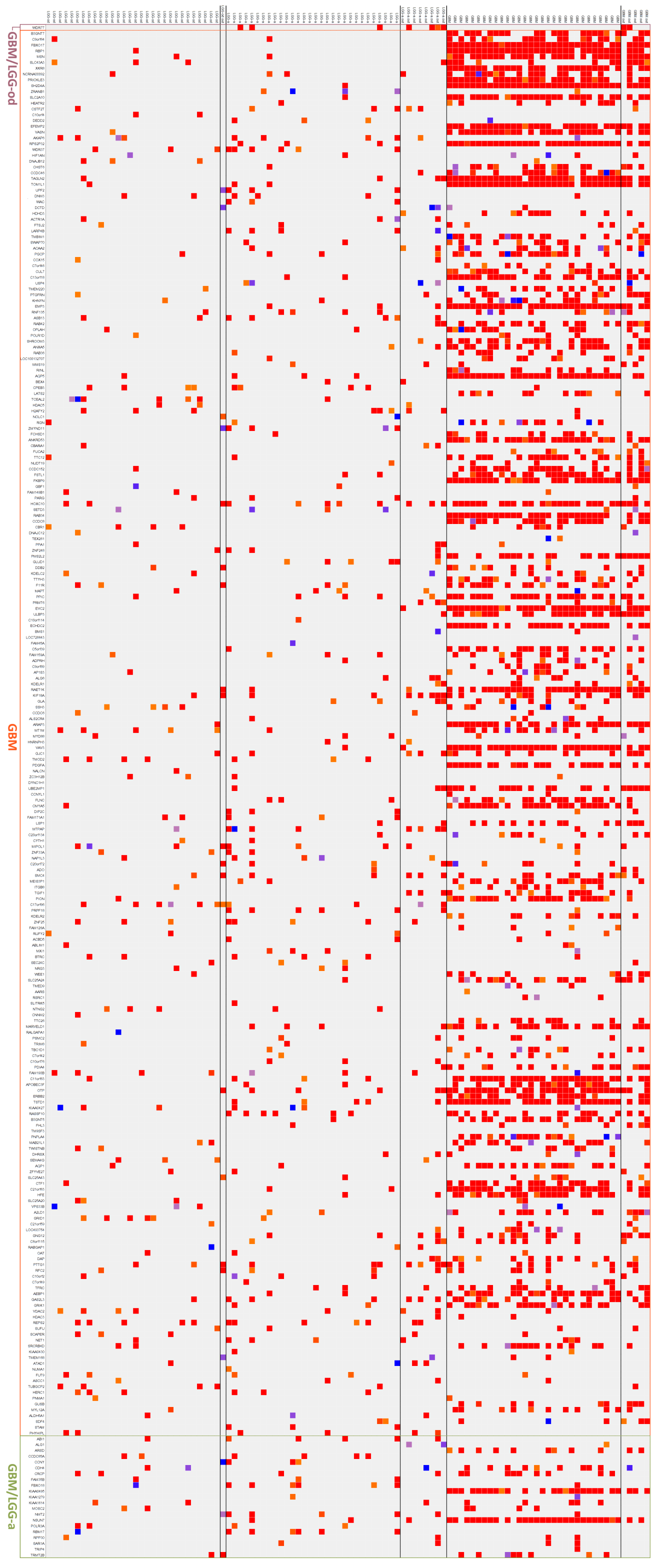


Figure S4: DDC plot for the features associated with GBM by the model from the RNA-seq dataset.

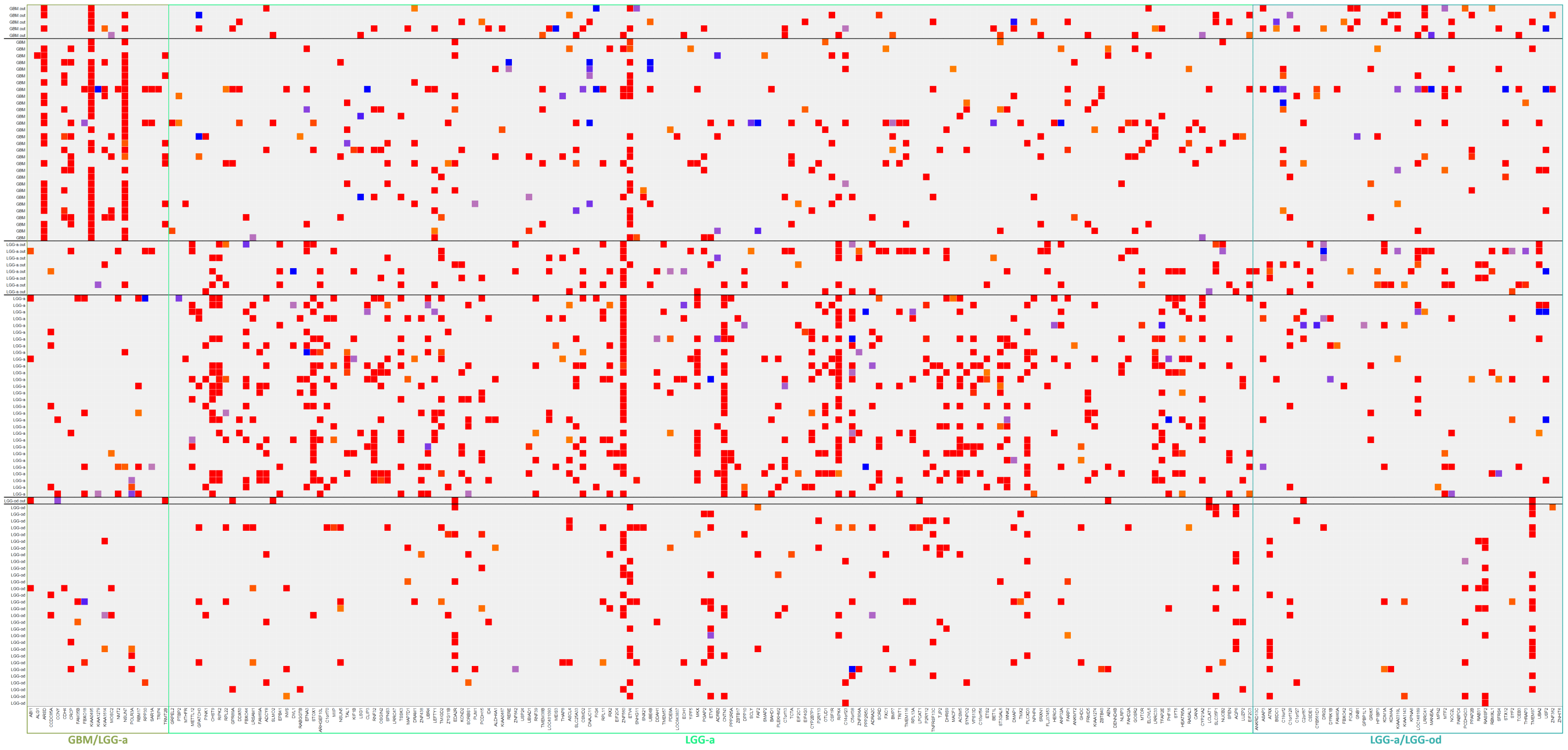


Figure S5: DDC plot for the features associated with LGG-a by the model from the RNA-seq dataset.

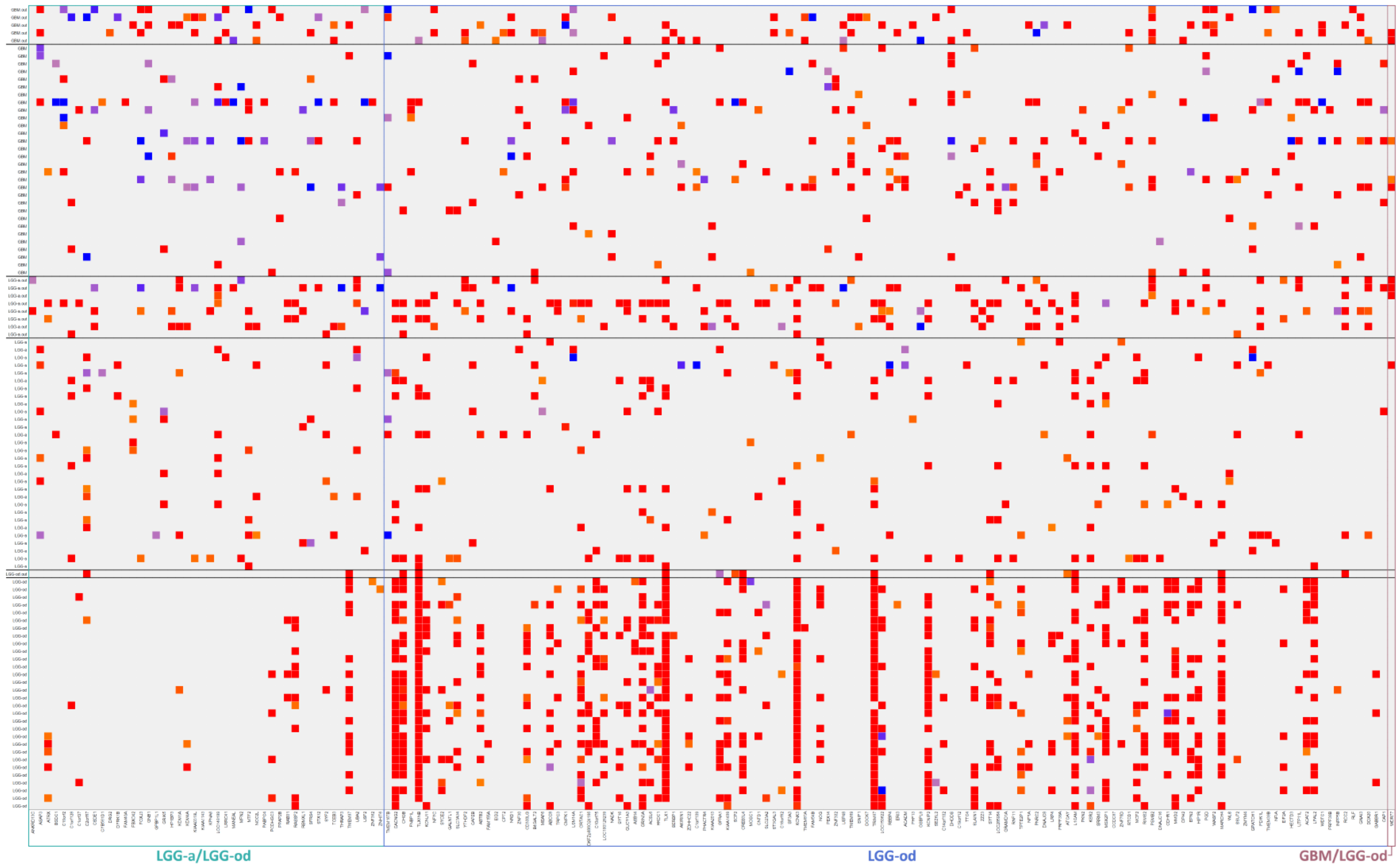


Figure S6: DDC plot for the features associated with LGG-od by the model from the RNA-seq dataset.

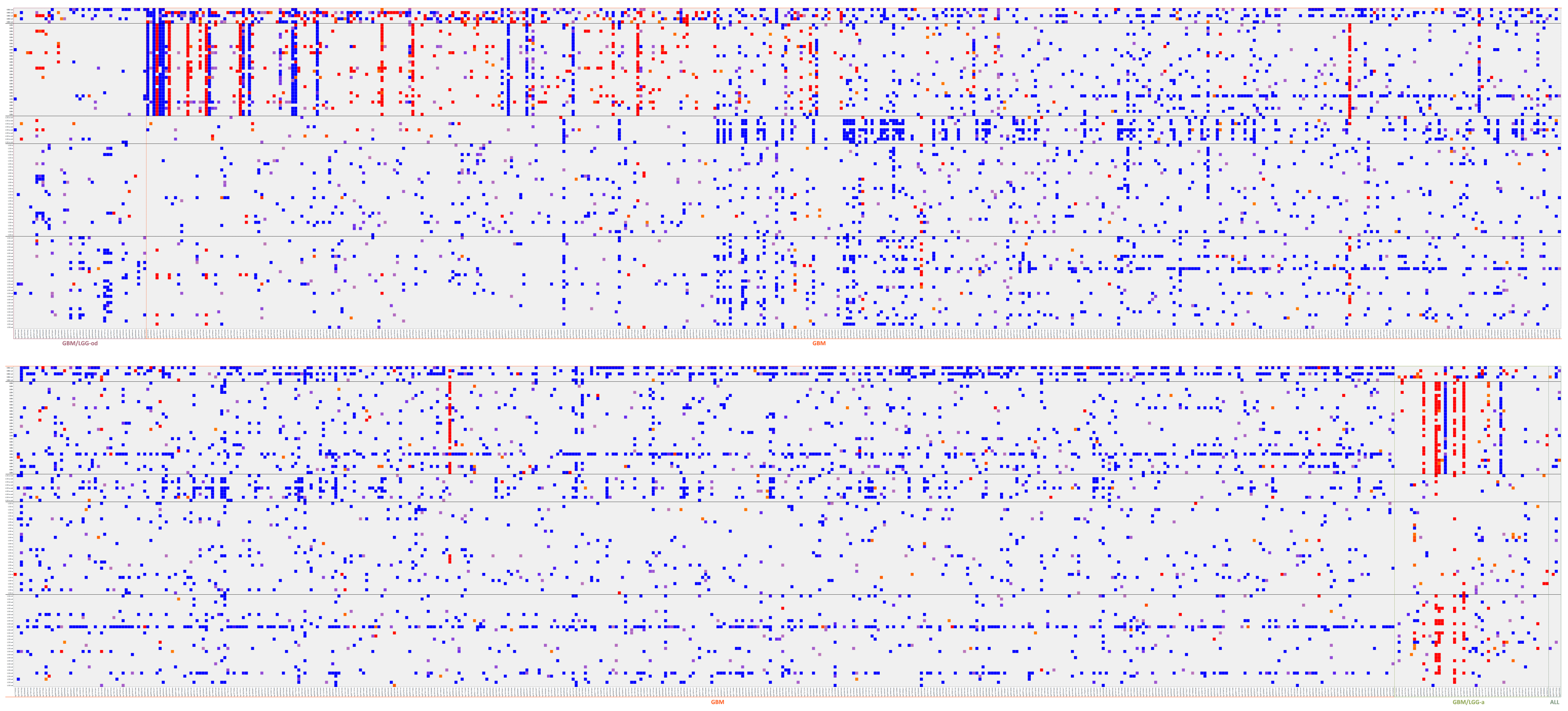


Figure S7: DDC plot for the features associated with GBM by the model from the DNA-meth dataset.

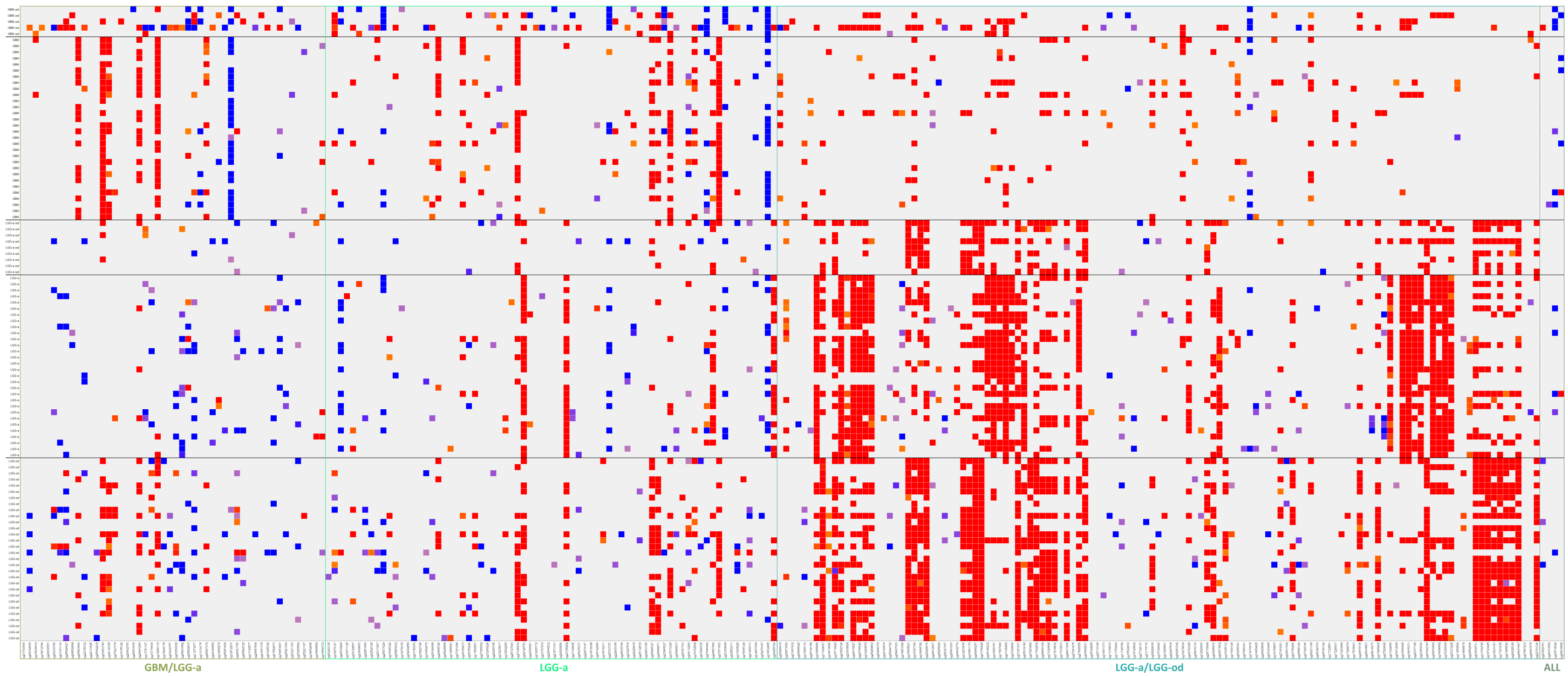


Figure S8: DDC plot for the features associated with LGG-a by the model from the DNA-meth dataset.

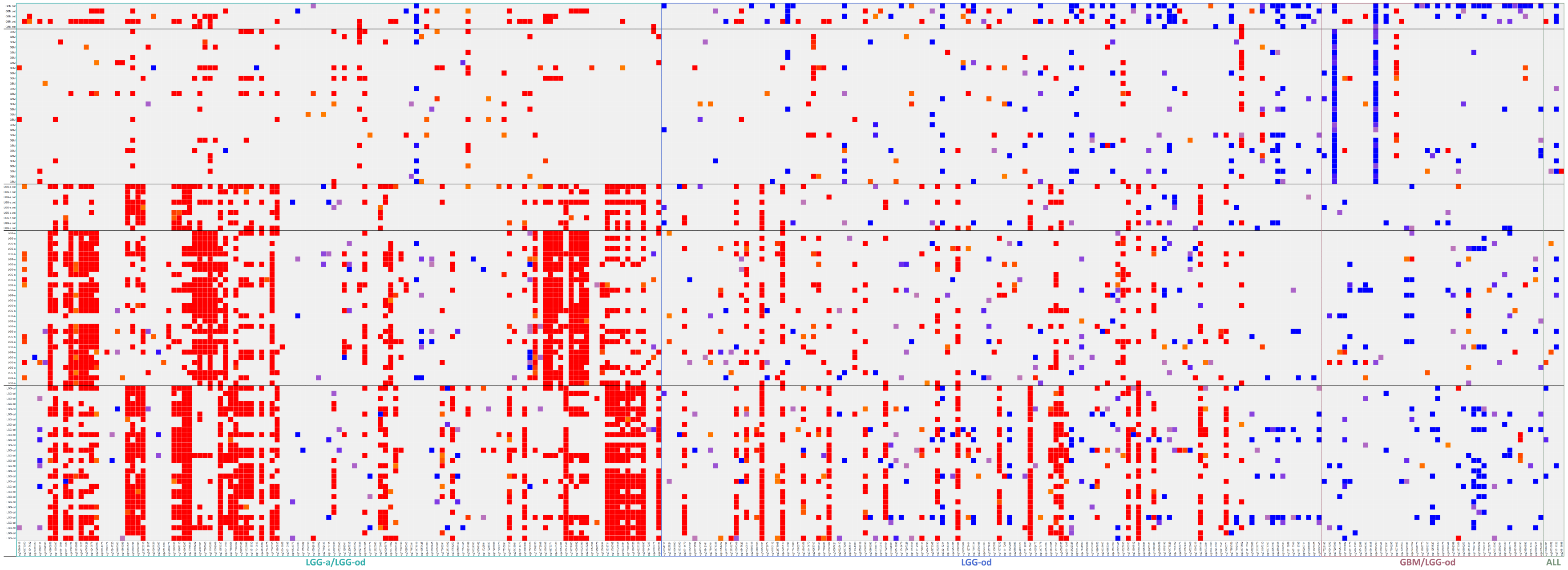


Figure S9: DDC plot for the features associated with LGG-od by the model from the DNA-meth dataset.

#### S3 Biomarkers selected by rSMLR models for 2021-WHO

Pages s12-s13 present Tables comprising the features selected by the rSMLR models, regarding 2021-WHO, as most relevant according to the respective absolute value of the respective coefficients' average. Due to the huge amount of selected features by each model (see Table **??**), only the top 20 features associated with each glioma type, for each model, was considered.

In Table S2, which presents the top 20 features obtained for the RNA-seq dataset regarding each glioma type, the numbers in the columns "GBM", "LGG-a" and "LGG-od" represent the position of the gene in the ordered list for GBM, LGG-a and LGG-od, respectively. Column "Knowledge" provides some insights about the reported links between the gene and tumoral diseases, if already reported ("N.R." stands for "not reported").

Table S3 presents the top 20 features obtained for the DNA-meth dataset regarding each glioma type. "$\mathrm{GBM_D}$", "LGG-$\mathrm{a_D}$" and "LGG-$\mathrm{od_D}$" columns show the number referring to the position of the methylation site in the ordered list for GBM, LGG-a and LGG-od, respectively. "$\mathrm{GBM_R}$", "LGG-$\mathrm{a_R}$" and "LGG-$\mathrm{od_R}$" columns recall the position of the gene linked to the methylation site in each ordered list, in case it is referred in Table S2.

Table S2: Top 20 of selected genes, for each glioma type.

| Gene | GBM | LGG-a | LGG-od | Knowledge | Ref. |
|---|---|---|---|---|---|
| *KIAA0495* | 1 | 68 | | Commonly overexpressed in oligodendroglial tumors | [1, 2] |
| *B3GNT7* | 2 | | | Highly expressed in primary astrocytic tumors | [3] |
| *C9orf4* | 3 | | | N.R. | – |
| *FBXO17* | 4 | | | Promotor of glioma development | [4] |
| *NSUN7* | 5 | 229 | | Associated with poor survival in liver cancer | [5] |
| *RBP1* | 6 | | | Nearly always hypermethylated in *IDH*-mutant tumors | [6] |
| *MSN* | 7 | | | miR-200c inhibits glioma progression via targeting *MSN* | [7] |
| *ARSD* | 8 | 50 | | Promotes glioma cells progression | [8] |
| *SLC43A3* | 9 | | | N.R. | – |
| *XKR8* | 10 | | | LGG progression and outcome | [9] |
| *NCRNA00092* | 11 | | | N.R. | – |
| *PRICKLE3* | 12 | | | N.R. | – |
| *KIAA1279* | 13 | 198 | | N.R. | – |
| *SH2D4A* | 14 | | | Exhibits tumor-suppressive functions | [10] |
| *ZRANB1* | 15 | | | Depressed expression in GBM | [11] |
| *CCNY* | 16 | 227 | | Inhibition of cell growth ability by suppression | [12, 13] |
| *SLC2A10* | 17 | | | Significantly highly expressed with poor prognosis GBM | [14] |
| *HEATR2* | 18 | | | N.R. | – |
| *CSTF2T* | 19 | | | Highly expressed in human melanoma cells | [15] |
| *C10orf4* | 20 | | | N.R. | – |
| *DRG2* | | 1 | 67 | Regulates immunity, imune signaling and metabolism | [16] |
| *TRIP4* | 58 | 2 | | High expression associated with poor overall survival | [17] |
| *BSDC1* | | 3 | 7 | N.R. | – |
| *GRPEL2* | | 4 | | redox regulator of mitochondria bioenergetics | [18] |
| *PTBP2* | | 5 | | Promotes proliferation and migration of glioma cell lines | [19] |
| *MTHFR* | | 6 | | c.677C¿T variant is a risk factor for survival in GBM | [20] |
| *METTL12* | | 7 | | Impact on mitochondria metabolism | [21] |
| *GPATCH3* | | 8 | | Involved in ocular and craniofacial development | [22] |
| *PINK1* | | 9 | | Negatively correlated with GBM growth and survival | [23] |
| *CHST9* | | 10 | | Downregulated in GBM | [24] |
| *FAM40A* | | 11 | 22 | Regulates cell contractility | [25] |
| *THRAP3* | | 12 | 125 | Depletion causes cellular hypersensitivity to DNA-damaging agents | [26] |
| *RIPK2* | | 13 | | Higher expression in TMZ-resistant glioma | [27] |
| *RPL22* | | 14 | | Decreased expression associated with multiple cancers | [28] |
| *GPR89A* | | 15 | | N.R. | – |
| *DDX50* | | 16 | | N.R. | – |
| *MOSC2* | 124 | 17 | | Progressive expression in colon tumors | [29] |
| *FBXO44* | | 18 | | Promotes DNA replication-coupled repetitive element silencing in cancer cells | [30] |
| *LRSAM1* | | 19 | | Positively correlated with survival in glioma | [31] |
| *FAM69A* | | 20 | | N.R. | – |
| *LRRC41* | | 106 | 1 | N.R. | – |
| *FBXO42* | | 95 | 2 | N.R. | – |
| *GPBP1L1* | | 97 | 3 | Related to breast cancer | [32] |
| *TMEM167B* | | | 4 | N.R. | – |
| *GNB1* | | 179 | 5 | Oncogene in cervical squamous cell carcinoma, retinoblastoma and lung cancer | [33] |
| *CACNG2* | | | 6 | N.R. | – |
| *HP1BP3* | | 174 | 8 | Overexpression enhances proliferation, self-renewal and TMZ resistance in GBM | [34] |
| *CHGB* | | | 9 | Positively correlated with overall survival in LGG | [35] |
| *SFRS4* | | 86 | 10 | N.R. | – |
| *FNBP1L* | | | 11 | N.R. | – |
| *TLX1NB* | | | 12 | Low expression associated with poor survival in LGG | [36] |
| *KCNJ11* | | | 13 | N.R. | – |
| *CSDE1* | | 48 | 14 | Poor prognosis in glioma | [37] |
| *ATRX* | | 25 | 15 | Regulator in IDH-mutant glioma | [38] |
| *NFYC* | | | 16 | Choroid Plexus Carcinoma oncogene | [39] |
| *SYCE2* | | | 17 | Potentiates DNA repair | [40] |
| *GALNTL1* | | | 18 | Down-regulated in breast cancer | [41] |
| *PABPC4* | | 139 | 19 | associated with triple-negative breast cancer | [42] |
| *TMEM97* | | 122 | 20 | Higher expression correlated with shorter survival time in glioma | [43] |

Table S3: Top 20 of selected methylation sites for each glioma type.

| Site | Linked gene | $GBM_D$ | $GBM_R$ | LGG-$a_D$ | LGG-$a_R$ | LGG-$od_D$ | LGG-$od_R$ |
|---|---|---|---|---|---|---|---|
| CG21816330 | *RAB34* | 1 | | | | | |
| CG19105122 | *SRRM3* | 2 | | 253 | | | |
| CG23056823 | *KIAA1614* | 3 | | | | | |
| CG12899157 | *TGIF1* | 4 | | | | | |
| CG03753331 | *DPP10* | 5 | | | | | |
| CG26146617 | *B3GNT7* | 6 | 2 | | | | |
| CG00916884 | *MT1M* | 7 | | | | | |
| CG05347878 | *NCRNA00092* | 8 | 11 | | | | |
| CG04075191 | *DPP10* | 9 | | | | | |
| CG07931631 | *C11orf63* | 10 | | | | | |
| CG01793449 | *KIAA0495* | 11 | 1 | 54 | 68 | | |
| CG14018731 | *NCRNA00092* | 12 | | | | | |
| CG20532370 | *RBP1* | 13 | 6 | | | | |
| CG03208951 | *EMP3* | 14 | | | | | |
| CG12604950 | *KIAA1614* | 15 | | | | | |
| CG18568589 | *ZNF560* | 16 | | 46 | | | |
| CG20129213 | *RIMS2* | 17 | | | | | |
| CG25042239 | *CUL7* | 18 | | 55 | | | |
| CG13952656 | *B3GNT7* | 19 | 2 | | | | |
| CG15555970 | *TGIF1* | 20 | | | | | |
| CG21195256 | *C14orf23* | | | 1 | | 1 | |
| CG24899806 | *KCND2* | | | 2 | | 3 | |
| CG18875371 | *C14orf23* | | | 3 | | 5 | |
| CG26816688 | *C7orf13* | 68 | | 4 | | 100 | |
| CG26333652 | *IRX2* | 133 | | 5 | | 95 | |
| CG23519022 | *CAPZB* | 243 | | 6 | | 135 | |
| CG03505995 | *C14orf23* | | | 7 | | 24 | |
| CG03909781 | *KIAA0495* | 292 | 1 | 8 | 68 | 229 | |
| CG21794428 | *LATS2* | | | 9 | | 2 | |
| CG12666279 | *DPP10* | 239 | | 10 | | | |
| CG16933181 | *CHST9* | | | 11 | 10 | 6 | |
| CG09874600 | *TRIP4* | | 58 | 12 | 2 | 84 | |
| CG27565555 | *BMP7* | | | 13 | | 138 | |
| CG23059304 | *C14orf23* | | | 14 | | 46 | |
| CG25104186 | *CSMD2* | | | 15 | | 68 | |
| CG07356745 | *FAM190B* | | | 16 | | 4 | |
| CG05346855 | *PPP2R5A* | | | 17 | | 7 | |
| CG13060114 | *TFAP2E* | | | 18 | | 155 | |
| CG19329389 | *TRIP4* | | 58 | 19 | 2 | 117 | |
| CG14006181 | *C14orf23* | | | 20 | | 61 | |
| CG15468521 | *TLX1NB* | | | 21 | | 8 | 12 |
| CG22928016 | *NRG3* | | | 38 | | 9 | |
| CG21895526 | *CHST9* | | | 29 | 10 | 10 | |
| CG13326686 | *SYCE2* | | | 22 | | 11 | 17 |
| CG19586576 | *GJC1* | | | 69 | | 12 | |
| CG07650391 | *LOC285954* | | | 24 | | 13 | |
| CG16723445 | *TGIF1* | | | 63 | | 14 | |
| CG26775866 | *PTTG1* | | | 39 | | 15 | |
| CG13709765 | *TGIF1* | | | 59 | | 16 | |
| CG22703724 | *ETS2* | | | 26 | | 17 | |
| CG08450106 | *SYCE2* | | | 27 | | 18 | 17 |
| CG18278638 | *SLC7A14* | | | 207 | | 19 | |
| CG01351037 | *PDE8A* | | | 117 | | 20 | |
